# Digital Exposure Notification Tools: A Global Landscape Analysis

**DOI:** 10.1101/2022.06.02.22275925

**Authors:** Camille Nebeker, Daniah Kareem, Aidan Yong, Rachel Kunowski, Mohsen Malekinejad, Eliah Aronoff-Spencer

## Abstract

**Background:** As the COVID-19 global pandemic continues, digital exposure notification systems are increasingly used to support traditional contact tracing and other preventive strategies. Likewise, a plethora of COVID-19 mobile apps have emerged.

**Objective:** To characterize the global landscape of pandemic related mobile apps, including digital exposure notification and contact tracing tools.

**Data Sources and Methods:** The following queries were entered into the Google search engine: “(*country name* COVID app) OR (COVID app *country name*) OR (COVID app *country name*+) OR (*country name*+ COVID app)”. The App Store, Google Play, and official government websites were then accessed to collect descriptive data for each app. Descriptive data were qualified and quantified using standard methods. COVID-19 Exposure Notification Systems (ENS) and non-Exposure Notification products were categorized and summarized to provide a global landscape review.

**Results:** Our search resulted in a global count of 224 COVID-19 mobile apps, in 127 countries. Of these 224 apps, 128 supported exposure notification, with 75 employing the Google Apple Exposure Notification (GAEN) app programming interface (API). Of the 75 apps using the GAEN API, 15 apps were developed using Exposure Notification Express, a GAEN turnkey solution. COVID-19 apps that did not include exposure notifications (n=96) focused on COVID-19 Self-Assessment (35·4%), COVID-19 Statistics and Information (32·3%), and COVID-19 Health Advice (29·2%).

**Conclusions:** The digital response to COVID-19 generated diverse and novel solutions to support non-pharmacologic public health interventions. More research is needed to evaluate the extent to which these services and strategies were useful in reducing viral transmission.

**Research in Context:** *Evidence before this study:* The COVID-19 pandemic created a role for technology to complement traditional contact tracing and mitigate the spread of disease. How countries responded with technology - specifically, how they utilized mobile apps to support public health was a focus of this research. The search process consisted of searching the Google Search Engine using queries “(*country name* COVID app) OR (COVID app *country name*) OR (COVID app *country name*+) OR (*country name*+ COVID app).” Apps that were found on the App Store, Google Play, and official government format that fit the pre-defined eligibility criteria were included in the Apps list considered in this search. Apps that did not match those criteria were excluded from the process. All 195 countries and associated COVID-19 apps were considered for inclusion if they were official COVID-19 apps adopted by the governments of those countries.

*Added value of this study:* Findings from this research contributes to the literature by providing a synthesis of how technology is being used to support public health authorities in reducing the spread of COVID-19. The results of this global landscape analysis of COVID 19 mobile apps, included both COVID-19 Exposure Notification Apps and non-Exposure Notification Apps.

*Implications of all the available evidence:* The findings of this research can provide the foundation for future studies to assess adoption rates and, subsequently identify app features that lead to increased adoption of both Exposure Notification and non-Exposure Notification Apps. Only when a product meets the needs of consumers will it be adopted and utilized, and in the case of exposure notification tools, save lives.

**Eligibility Criteria:** The study eligibility criteria included any COVID-19 mobile applications and COVID-19 Exposure Notification (EN) systems that were downloadable or activated on a mobile device. The search process involved using queries “(*country name* COVID app) OR (COVID app *country name*), OR (COVID app *country name*+) OR (*country name*+ COVID app)” in Google search engine to identify COVID-19 apps in each country. The data included active and inactive apps discovered during the data collection period of June 2021 - July 2021.

## Introduction

On March 11, 2020, SARS-CoV-2, known as COVID-19, was officially declared a global pandemic by the World Health Organization (WHO)^1^. As of March 2022, over 450 million confirmed cases of COVID-19 and 6 million deaths have been reported^2^ Efforts to reduce infection spread have led to myriad digital exposure notification tools to support traditional public health contact tracing strategies.

The goal of contact tracing is to identify those who have been exposed to a particular communicable disease and isolate them before they further spread the disease. This disease mitigation method dates back at least to the Middle Ages when red crosses were painted atop homes sick with the Plague and ships routinely quarantined for 40 days (Italian (venetian) *quarantine* ‘forty days’)^3,4^. Traditional contact tracing works by identifying and contacting persons exposed to the infected individuals to see who they have been in contact with and so forth. Exposed contacts are notified and must isolate if they test positive, averting an otherwise new transmission chain. These methods have significant draw backs. Individuals may be reticent to engage with contact tracers, not be forthcoming with contacts, and may not remember their past contacts^5,6^. Further, the resources to conduct manual contact tracing can be rapidly overcome as case rates rise, and coordination across jurisdictions can be nearly impossible. Today, digital solutions can supplement and work in a synergistic manner to address many of these shortcomings.

Exposure Notification (EN) systems can play an important role in managing the spread of COVID-19 and other infectious diseases^7.^ Exposure Notification and traditional contact tracing have similar goals – that being to notify people who may have been exposed to the virus^8^. The ability to leverage technology to detect and communicate exposure can complement traditional methods and save lives^7^. A specific instance where digital EN can compensate for the limitations of traditional contact tracing is in instances where one is in proximity with strangers whom you would be unable to identify and therefore be unable to notify of exposure with the traditional contact tracing method^9^. Using EN technology, even strangers can be notified of their potential exposure and therefore help mitigate the spread of the virus.

Digital EN is designed to alert users of potential exposure to COVID-19, as well as to provide information and referral to testing and other forms of support. Bluetooth technology is one strategy used to support digital EN where anonymous “keys” are exchanged based on users distance and duration of exposure. Some apps use Global Positioning System (GPS) tracking and QR code status as digital EN strategies, GAEN API used a decentralized data collection system and majority of apps relied on Bluetooth technology for EN due to privacy protections^8,10^. In a partnership between Google and Apple, the Google Apple Exposure Notification System (GAEN) was rapidly developed in April 2020, which followed the PACT Protocol (Privacy Automated Contact Tracing) designed to preserve privacy in exposure detection methods^11^. GAEN was created for public health authorities (PHAs) to develop their own Exposure Notifications app using the GAEN API. Exposure Notifications Express (ENX) was a later development using GAEN. With ENX, PHAs provide Google and Apple with a configuration file that contains instructions, content, messaging, and risk parameters, which uses the GAEN API to generate a custom exposure notification app.

## Methods

The purpose of this global landscape analysis was to document and characterize COVID-19 mobile apps, including Exposure Notification (EN) technologies.

### Eligibility Criteria

The study eligibility criteria included any COVID-19 mobile app and COVID-19 Exposure Notification (EN) systems that were downloadable or activated on a mobile device. The search process involved using queries “(*country name* COVID app) OR (COVID app *country name*) OR (COVID app *country name*+) OR (*country name*+ COVID app)” in the Google search engine to identify COVID-19 apps in each country. The data included active and inactive apps discovered during the data collection period of June 2021 - July 2021.

### COVID-19 App Selection Strategy

The process for screening COVID-19 mobile apps involved first identifying if the COVID-19 app was a mobile app, which was defined as being able to download on a mobile device. Three co-authors (DK, AY, RK) divided a list of 195 countries into thirds. Each co-author then identified all COVID-19 apps that existed or previously existed in those countries. Our review includes both COVID-19 Exposure Notification (EN) apps and non-Exposure COVID-19 apps (non EN).

### Data Collection Process

Identified apps were entered into a database by country where the app was deployed along with descriptors of the app functionalities. When app descriptions in news articles, app websites, Google Play Store, or App Store were not available in English, Google Translate was used for translation.

The data collection process was divided into two phases. Phase 1 consisted of identifying every COVID-19 related mobile app using the above search method. For each resulting app, descriptive data were entered into a spreadsheet. The data included whether the: 1- app was an exposure notification app and, if yes, whether the API was GAEN, 2- product use was voluntary or mandatory in the country, and 3- product used privacy-first messaging on the official app website, Google Play Store, and/or App Store. In addition, the dataset included data management and design considerations.

Phase 2 involved separating apps into two categories: 1- Exposure Notifications apps (EN) and 2- Non Exposure Notification apps (non EN). EN apps were further categorized as using GAEN and/or Exposure Notifications Express (ENX). For each COVID-19 app, the features were documented, and descriptive statistics applied.

Assumptions made in the process were that the information given on the Google Play store, App Store, and articles/websites discussing the app accurately described all app features. Table 1 below lists the app features and intended functionality.

**Table 1.**
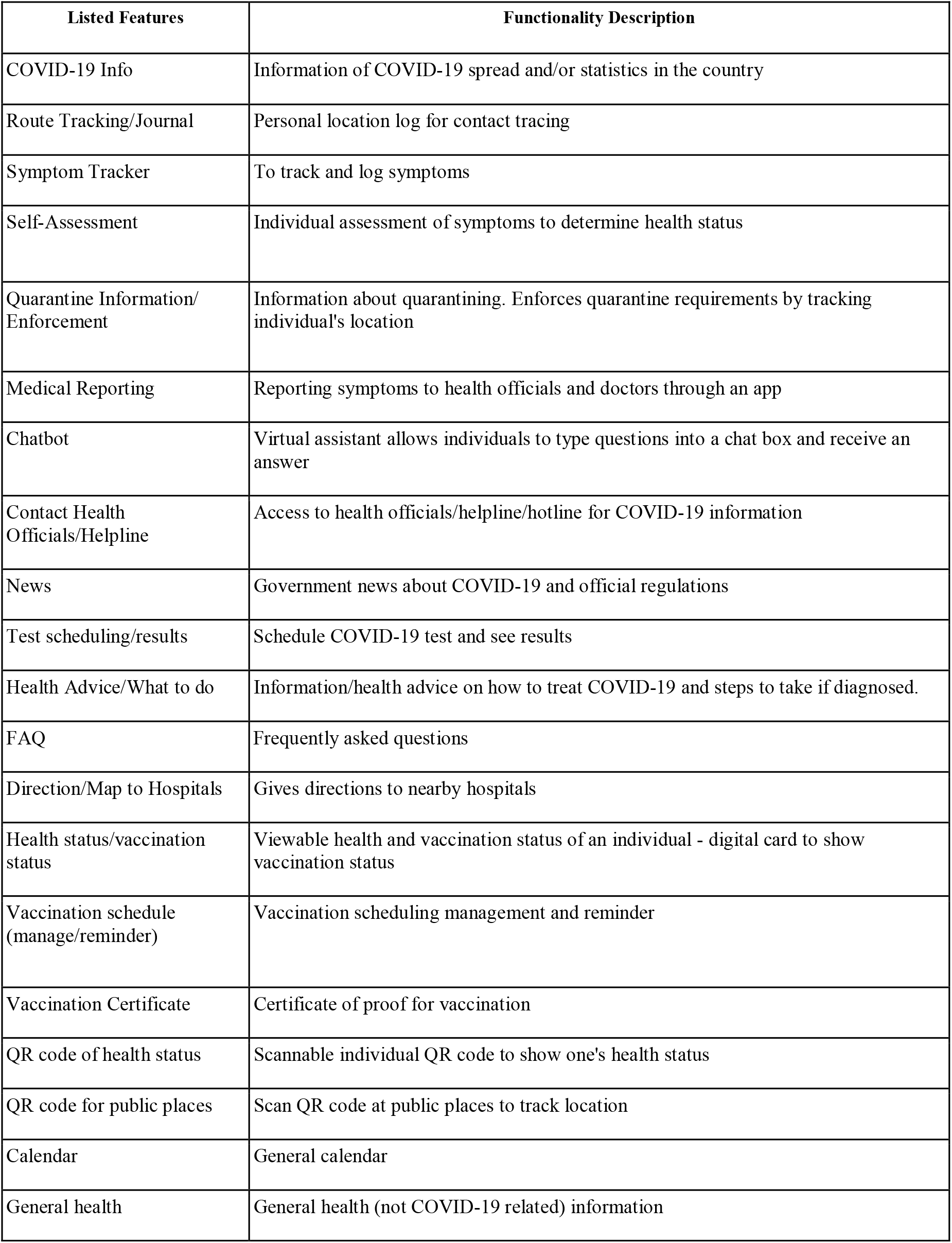
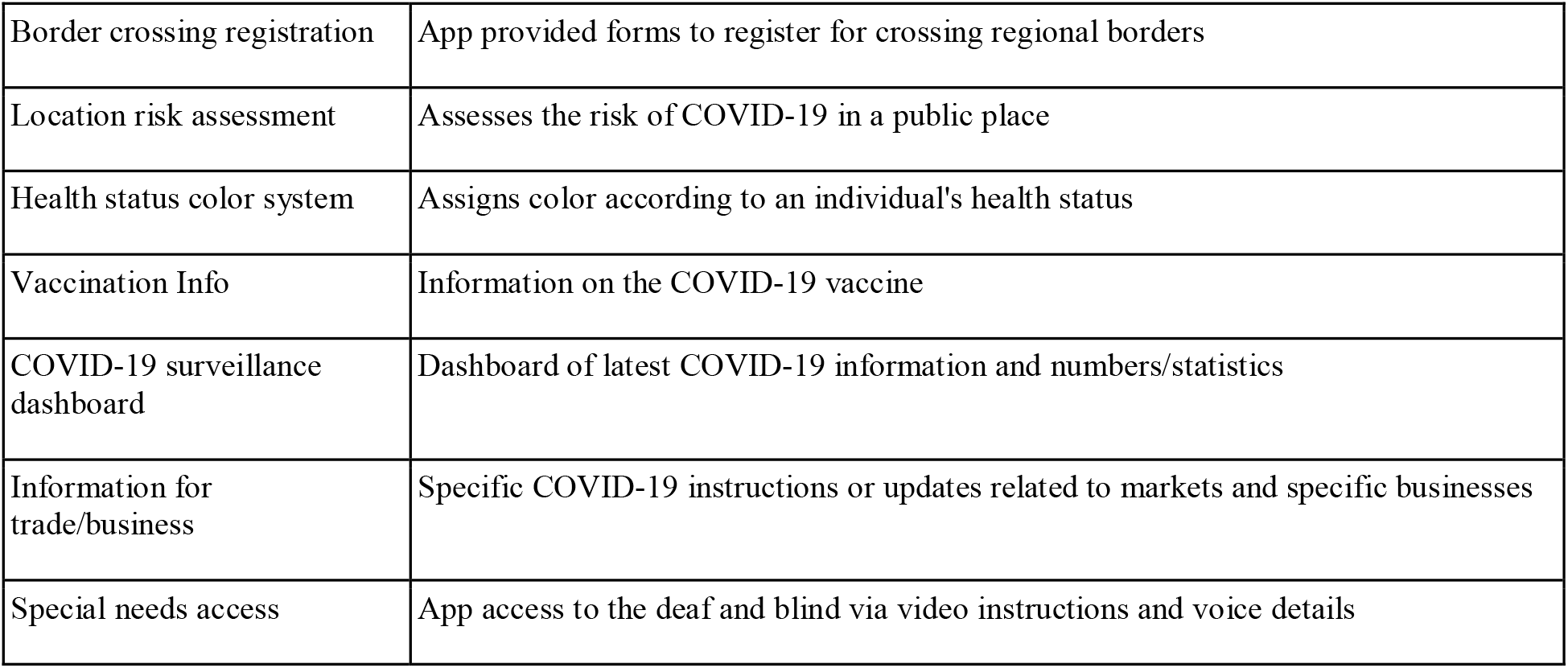
COVID-19 apps features and a description of intended functionality.

A second review involved authors cross checking the data collection and analysis process. The app search process involved three researchers inputting the same keywords into the same search engine on three different laptops, (*country name* COVID App). The researchers checked for consistency in coding by searching for apps within the same 10 countries independently and then met to discuss the codes used to describe features and develop code definitions. Upon agreement of the coding schema, the search for global apps was divided among the three co-authors, with each responsible for searching countries within the assigned segment and documenting all apps and functionalities. To consolidate the coding schema, a codebook was created to define each documented functionality. a second review involved authors cross-checking a different section of the country list than they had previously reviewed. All discrepancies were then addressed in a meeting between the three researchers and missing eligible apps were added to apps list.

## Results

The results describe characteristics and functionality of 224 COVID-19 mobile apps available globally. The data were drawn from 195 countries, including the United States and each of its 50 states.

### Global Digital EN Landscape

Figure 1 shows a color-coded geographic map of all the countries/regions using COVID-19 Exposure Notification Apps by API.^*1^

**Figure 1.**
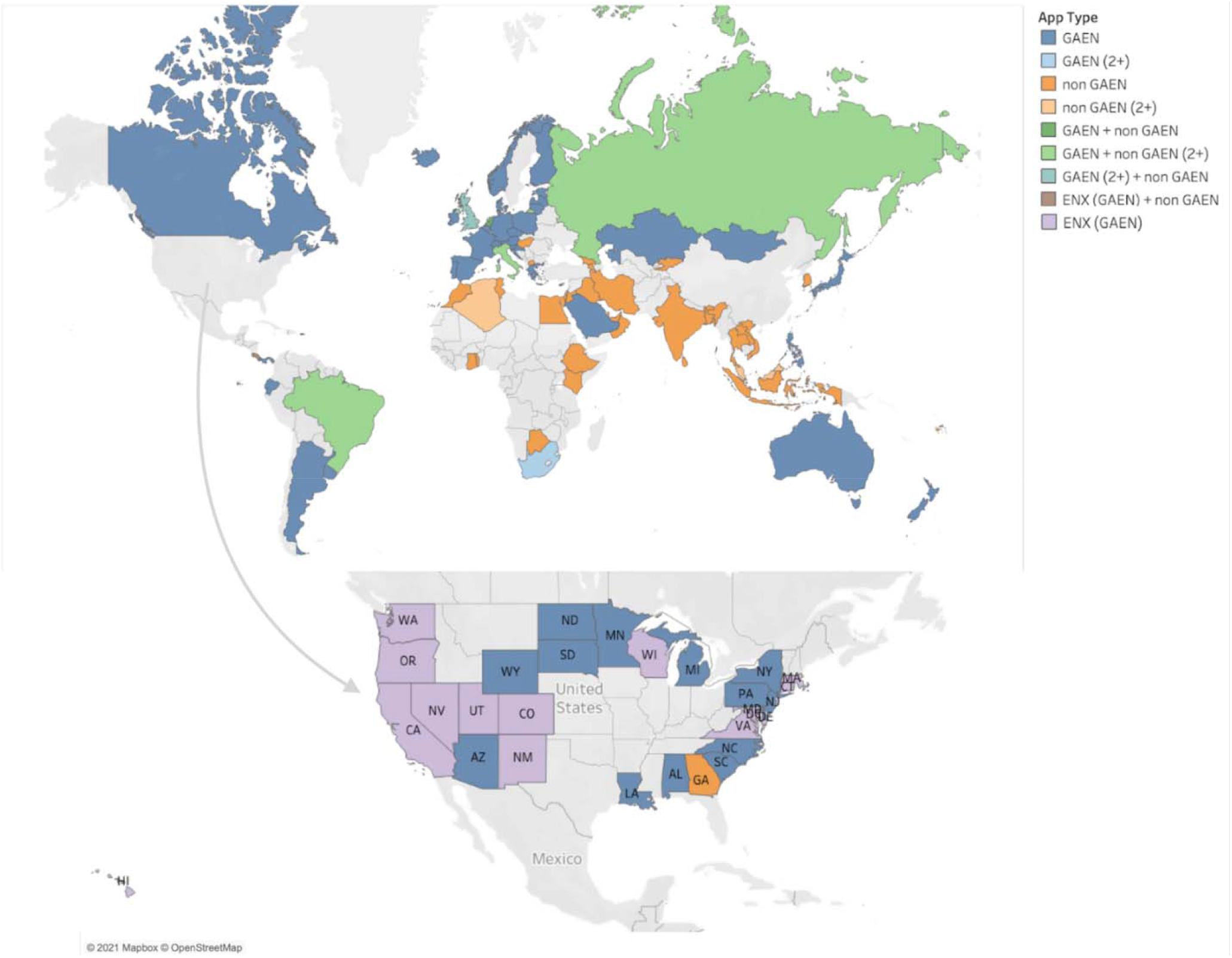
Global map of regions categorized by Exposure Notification (EN) App API.

#### Voluntary vs Mandated

Of the 224 COVID-19 apps, 150 apps (67·0%) were available for voluntary use where the user may choose to opt-in or out at their convenience. Conversely, 15 apps (6·7%) mandated user adoption and the remaining 59 apps were unclassified (26·3%). Mandates were enforced by either the government directly (n=7) where there were fines and consequences for not using the apps, or by the institution (e.g., school, airports, etc.) (n=5), or by workplace (n=3).

#### EN vs Non EN

Of the total, 128 apps (57·1%) were EN apps and 96 (42·9%) were non EN apps with 75 of the EN apps (58·6%) built with GAEN. Within the 75 GAEN apps, 15 apps (10·4% of all EN apps) used the Exposure Notification Express (ENX) system. The remaining 53 apps (41·4%) used an API other than GAEN (see figure 2).

**Figure 2.**
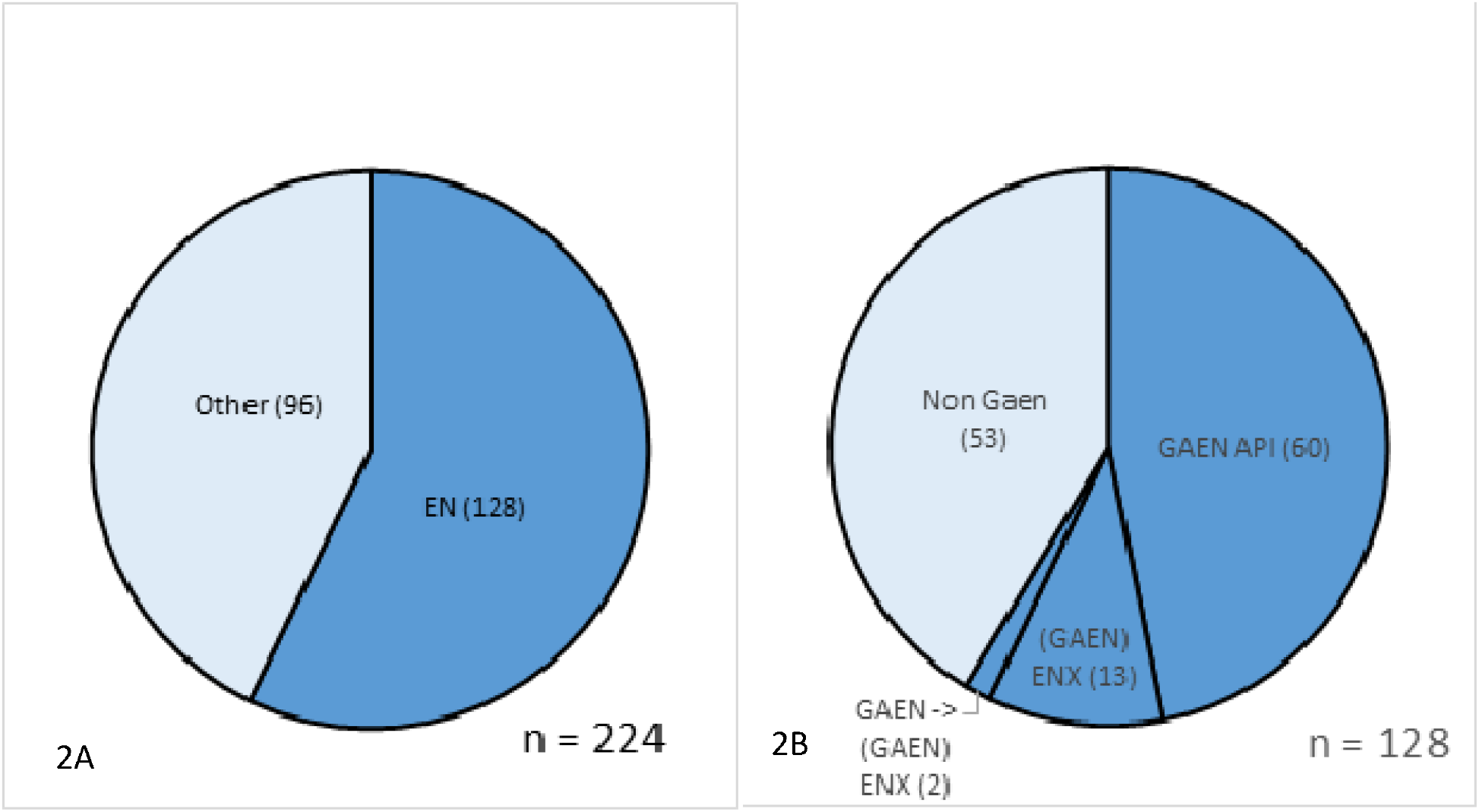
EN App Features: Figure 2a depicts the numbers of exposure notification and non-exposure notification apps and Figure 2b depicts the type of API technology used.

### EN Apps Functions

The primary function of the 128 Exposure Notification Apps is to support communication of exposure to COVID-19. Most apps included additional features including the availability of COVID-19 information (n = 22, 17·2% of apps), self-assessment guidance (n = 21, 16·4% of apps), health advice (n = 20, 15·6% of apps), test scheduling and test results (n =13, 10·2% of apps) and contact information for health officials (n = 12, 9·4%) (see figure 3a below).

**Figure 3.**
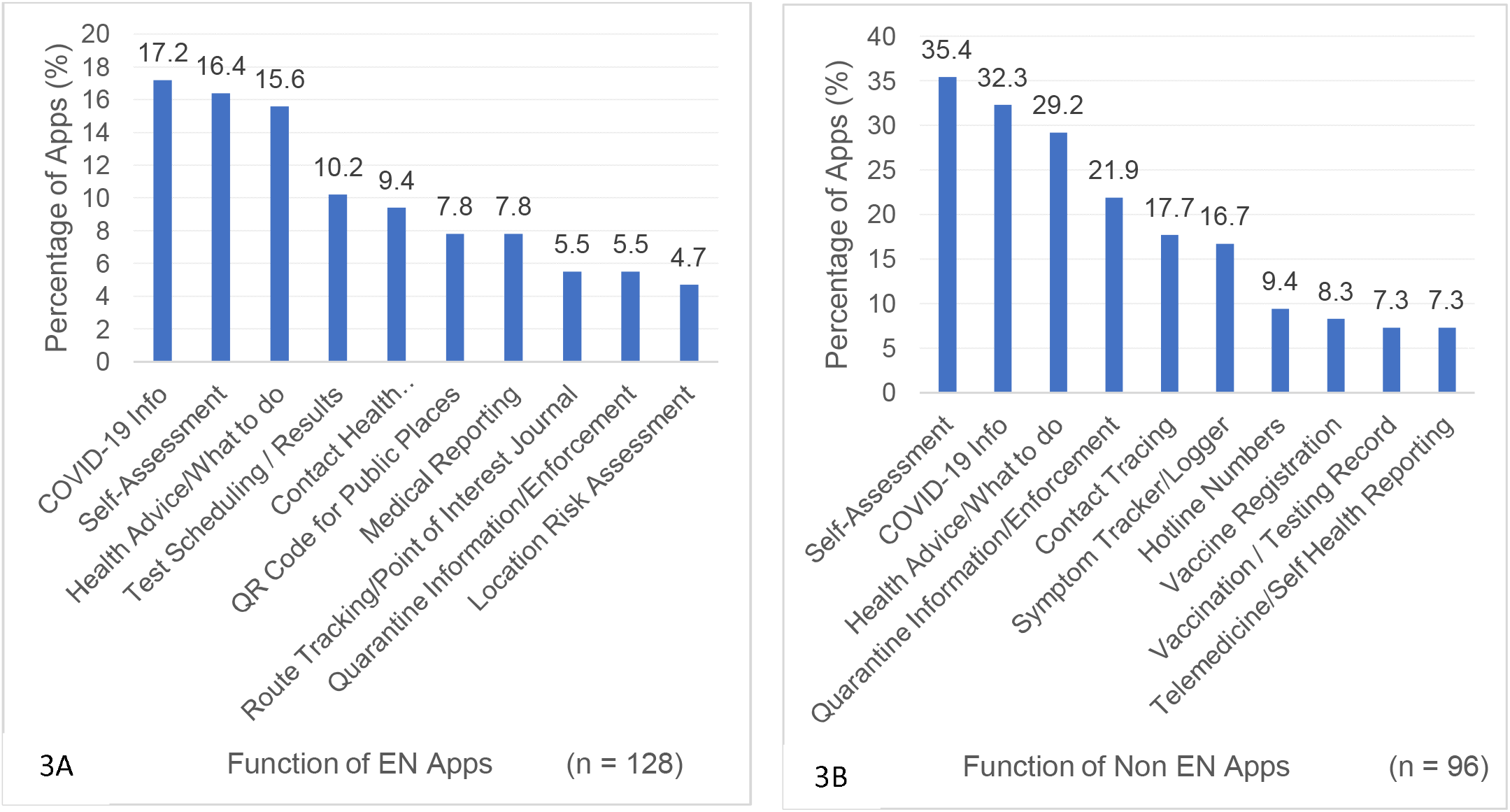
App Functions: Figure 3a consists of the 10 most prevalent functions across 128 Exposure Notification apps. Figure 3b depicts the 10 most prevalent functions across the 96 non exposure notification apps.

### Non EN Apps Features

The features in the 96 non EN apps are similar to the EN apps except they do not include exposure notification. The top 5 features found in non EN apps were self-assessment guidance (n=34, 35·4% of apps), COVID-19 information and statistics (n=31, 32·3% of apps), health advice (n=28, 29·2% of apps), quarantine information and enforcement (n=21, 21·9% of apps), and contact tracing (n=17, 17·7%) (see figure 3b above). Contact tracing in non EN apps differed from contact tracing in EN apps. In the non EN Apps, the app offered an option to report to PHAs if the individual had tested positive for COVID-19. The PHA would then handles the contact tracing process, instead of it occurring digitally like in the EN apps.

### ENX Apps Features

As noted previously, Exposure Notification Express (ENX) apps use the GAEN foundational platform but can be tailored to fit the needs of a specific state or organization, as such, the features are like EN. For example, Hawaii’s AlohaSafe Alert, Nevada’s Covid Trace Nevada, and Virginia’s COVIDWISE apps maintain unique app features because they deployed an original GAEN exposure notification app and then later adopted the ENX turnkey solution, which maintained their original app features.

There were 15 ENX apps identified with all 15 being used in the US. Figure 4 illustrates the percentage of apps containing the specified function. The top 5 features found in ENX apps include Upload Your COVID-19 Test Results (n=15, 100·0% of apps), Anonymously Alert Others of Exposure (n=15, 100·0% of apps), Review Your COVID-19 Exposure Status (n=15, 100·0% of apps), Learn about EN (n=13, 14·3% of apps), and Share the Word/Invite Others (n=13, 14·3% of apps).

**Figure 4.**
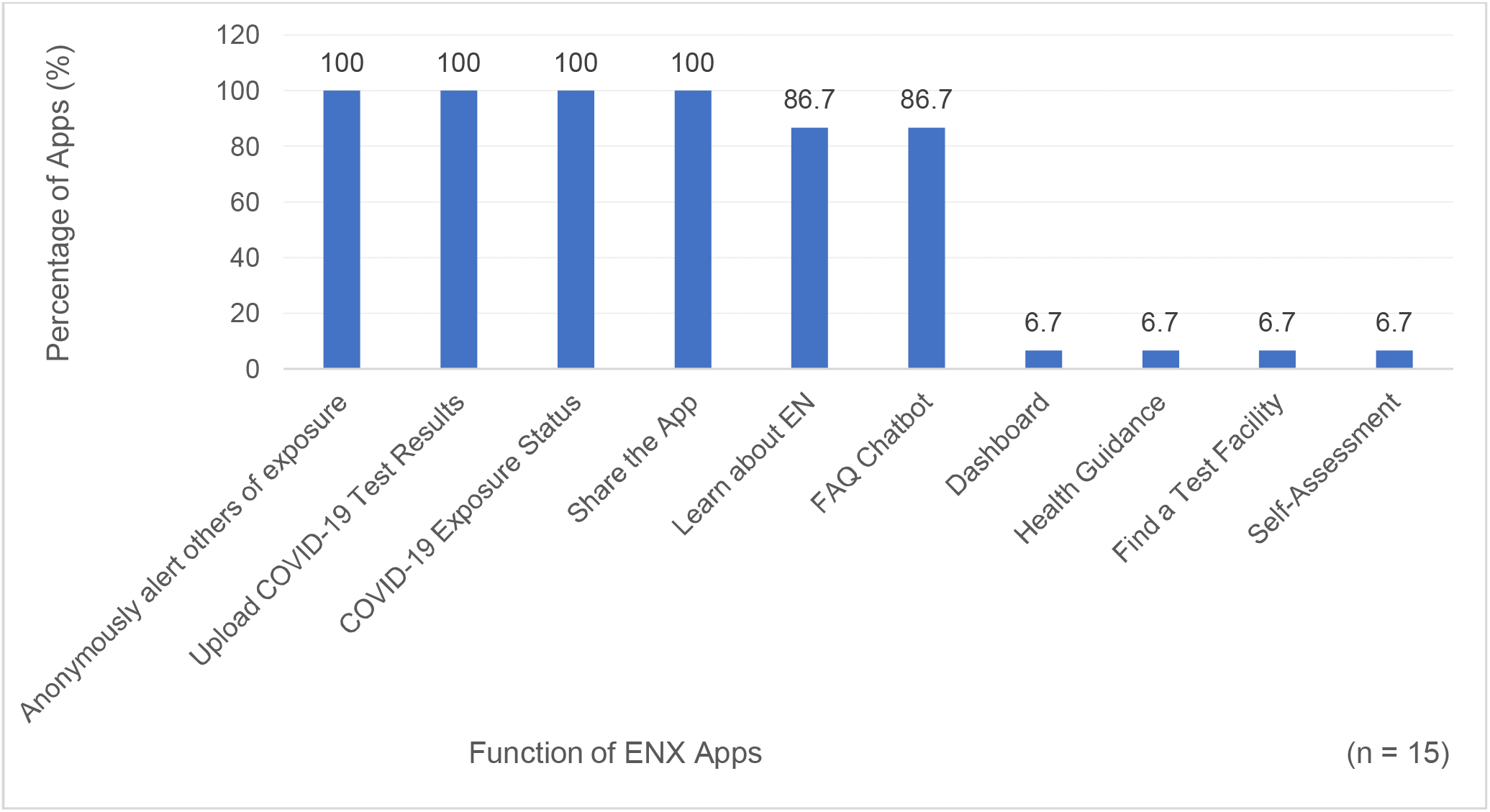
ENX Apps Functions: Figure 4 illustrates the most prevalent functions present across 15 ENX apps.

### GAEN and Non-GAEN Apps Features

Figure 5a shows the prevalence of functions in apps using the GAEN API (n=75). The most common features include: COVID-19 Info (n = 13, 17·3% of apps), Self-Assessment (n = 12, 16·0% of apps), Health Advice/What to Do (n = 11, 14·7% of apps), Contact Health Officials/Helpline, (n = 6, 8·0% of apps), Medical Reporting (n = 6, 8·0% of apps), and Test Scheduling/Results (n = 6, 8·0% of apps).

**Figure 5.**
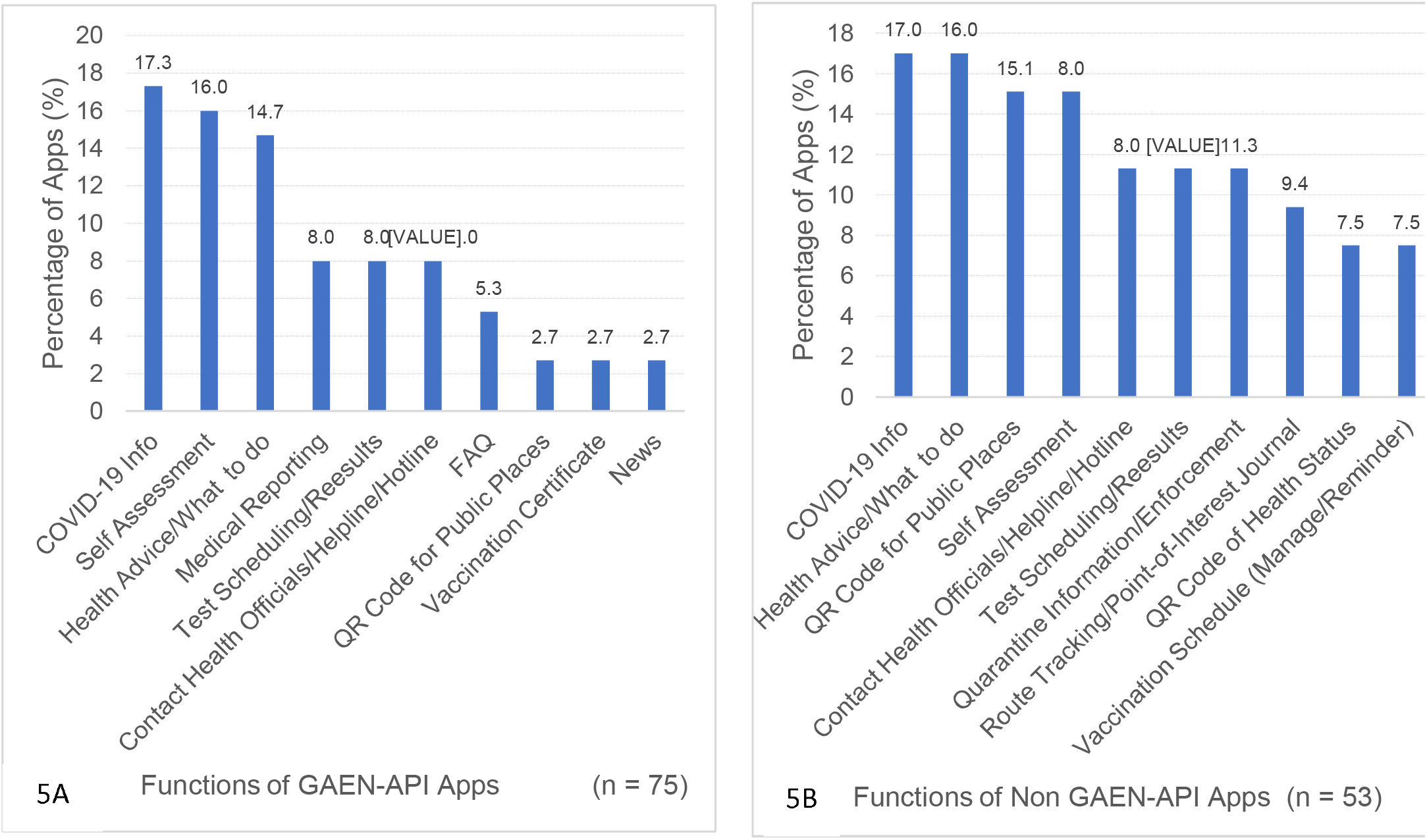
Functions in EN apps by API. Figure 5a functions in GAEN apps per total count of functions found in GAEN apps. Figure 5b functions in non-GAEN EN apps per total count of functions found in non-GAEN EN apps.

Apps not using the GAEN API (non-GAEN) make up 41.4% (n=53) of COVID-19 Exposure Notification apps. Figure 5b identifies the most prevalent features within the EN apps that use an API other than GAEN. The following are the five most prevalent functions found in non-GAEN apps: COVID-19 information (n=9, 17.0% of apps), Health Advice/What to Do (n=9, 17.0% of apps), Self-Assessment (n=8, 15.1% of apps), QR Code for public places (n=8, 15.1% of apps), and Contact Health Officials (n=6, 11.3% of apps). Compared with GAEN apps, they share the same top three most prevalent functions: COVID-19 information, Self-Assessment, and Health Advice/What to Do. Regardless of whether the apps used GAEN or non GAEN API, EN apps share many of the same core features. Non GAEN apps had additional features, including: QR codes for public places (15.1% of non GAEN apps), and quarantine enforcement/information (11.3% of non GAEN apps), which were not present in GAEN apps. The difference in functions can be attributed to the different main purpose of GAEN based apps versus non GAEN apps and the needs and features prioritized by where they were adopted.

#### Privacy

Designers of the apps and governments seemed to be aware of privacy being a concern based on the way some apps were designed and advertised with users’ privacy in mind. Our findings show that 56 Apps, 25% of the total Apps, had some sort of privacy messaging or claimed to value privacy, whereas 44% of Exposure Notification apps included a form of privacy first messaging. For example, in Jordan’s App store description for its EN app, AMAN, the app description states:

“Your privacy matters; it is as important as your health. Therefore, AMAN only stores data on your phone and does not request any personal information or data that could lead to your identification or to breaching your privacy.”

#### Data Management

Thirty-three of the 224 apps (14·7%) had centralized data collection where data collected by the app were managed through a centralized government server or institutional data storage system. Ninety-three (41·5%) COVID apps used a decentralized data management system with data saved on the phone that had the app and was not transferred or saved somewhere else.

#### US vs Global COVID Apps

The US had a total of 33 COVID-19 Apps being used across the 50 states and Washington D.C.; however, no app was available nationally. Twenty-seven of these (81.8%) were EN apps. Each state had its own unique app and all the US apps that were made by the state and, not for a work or institution, were opt-in. The US has the highest number of COVID-19 apps compared to other countries. Covid apps that were associated with the state universities, but not adopted statewide, were still included in the eligibility criteria.

#### Context

While COVID-19 exposure notification apps are intended to limit the spread of infection, the existence of a COVID-19 EN app does not necessarily define the density of COVID-19 infections in the countries that used them. The countries with the lowest prevalence of COVID-19 did not necessarily have COVID-19 apps helping them. Moreover, limited data exists regarding adoption as well as effectiveness in mitigating spread due to the recentness of COVID-19, the novelty of exposure notification technology and privacy protections.

## DISCUSSION

There was an unspoken consensus across the world that technology can be a useful tool in mitigating the spread of the COVID-19 pandemic, which was demonstrated by advances in the development and demand of novel health technologies, supporting remote monitoring, telemedicine, and digital public health interventions such as EN^13^. In this light, it is informative to compare how managing the spread of COVID differed from past pandemics. To our knowledge, this is the first pandemic to utilize technology to track exposure and augment traditional public health contact tracing. A recent study of COVID-19 technology reported the types of technologies used and adopted in digital exposure notification, including which countries adopted the various technologies and related challenges and concerns with each^8^. Our research adds to this literature by providing the first comprehensive global analysis of all COVID-19 mobile apps, their APIs, and functions provided by those apps for both Exposure Notification Apps (EN) and non-Exposure Notification Apps (non EN).

Digital contact tracing and exposure notification facilitate access, speed of information exchange, and can be scaled in a manner that is not achievable through traditional contact tracing methods. Another benefit is the infrastructure, resources and people needed to support traditional contact tracing may be quickly overwhelmed and digital solutions may allow human resources to be more effectively applied. While digital contact tracing and exposure notification sound the same, they have some fundamental differences. Digital contact tracing in COVID-19 apps is a new kind of technology where infected persons use their phones to anonymously send an exposure notification to responsible health officials who inform other potentially exposed parties. Exposure notification technologies may use WiFi, Global Positioning System (GPS), QR Codes, or Bluetooth technology to record devices in proximity^8^. Once a person has been exposed or tests positive, they can notify the App by the click of a button. The App will then send an “exposure notification” message to the devices that have been in proximity. This transfer of proximity information can happen anonymously, when using stored Bluetooth data. Among the 36 countries that had more than one COVID App, these apps were either complementary, where some apps operated as exposure notification apps, and the other for COVID stats and updates, or they were for specific provinces and institutions within the country. Eight countries had more than one EN app, which could have resulted from competition or specificity with a region. Some countries had both their own Exposure Notification app and a GAEN app. It would be interesting to see, as data becomes more available, which API had greater usage and what kind of app was more effective in mitigating the spread of COVID-19. In addition, identifying whether the use of Bluetooth and privacy protecting programming led to greater adoption. The tradeoff of privacy protection is the ability to know more about system performance and disease spread. Knowing exposure location could be a game changer with respect to deploying mitigation strategies in specific regions, yet may present unintended consequences. At this time, data are being collected to identify whether those using EN in the United States would use the technology if GPS were a feature of the EN, which, if found useful, can be integrated into updates.

Distinguishing COVID-19 Exposure Notification (EN) apps by their API can be useful in determining what makes an app adoptable and successful in reaching its audience. Although within this study there were limitations in measuring the adoption and usage of apps, doing so would offer insight as to what makes an app attractive and achieve higher rates of adoption over other apps. Another limitation is the time frame where the data collection process took place, as the landscape may shift significantly since then. The measurement of an exposure notification system’s effectiveness in limiting the spread of infectious diseases, which was not conducted within this study due to various limitations, would also be of great significance for the development of future EN apps.

### Privacy Concern

A Swiss Survey found that only 1 out of 3 citizens downloaded their EN app, SwissCovid, for which 24% of respondents reported privacy is a concern^14^. GAEN technology was specifically created with “user privacy and security central to the design.”^15^ Notably, most apps were designed within a few months of COVID-19 being declared as a pandemic on March 11th, 2020^1^, demonstrating the wide acceptance of COVID-19 apps as a valuable resource. This topic of privacy is of great ethical importance that needs to be continuously monitored.

### Functions

Our data reveal a global consensus that technology can significantly contribute to the mitigation of COVID-19, whether through exposure notification, general information, or any other functions noted. The three most prevalent functions in both EN and Non EN apps were COVID-19 info, self-assessment, and health advice, which demonstrates a majority COVID-19 apps are used to inform the public regardless of EN capabilities. All EN apps, as expected, had the primary function of notification; therefore, secondary functions were not likely prioritized during product development (e.g., a main function of providing COVID-19 information was 16% in EN and 33% in Non EN apps). Given apps maybe more acceptable with added features, developers may want to consider providing more app functions in future iterations.

### Limitations and Future Directions

Due to the fast-paced development and deployment of COVID-19 apps and the uncertainties of a global pandemic, there was limited research available about app adoption or effectiveness. While not perfect, the findings from this study provides insight on technology usage in this current pandemic and can be leveraged to learn shortcomings of COVID-19 apps for future pandemics and epidemics.

Possible bias or inaccuracy may have occurred when apps found were only accessible in their respective country’s native language. Google Translate was used to interpret the official app pages and the graphic images describing the function of the apps in the App Store and Google Play. In addition, personalized algorithms could influence which apps the Google Search engine pushed prioritized on the results page. The landscape of COVID-19 apps additionally may shift as time passes on.

Within this study, apps using the GAEN API make up 56·0% of COVID-19 exposure notification apps globally. Analyzing the adoption rates and impact of apps using the GAEN API compared to apps using other systems would be beneficial for developing future technology with higher adoption and effectiveness. This study did not pursue data on adoption rates due to lack of current availability. However, future research should pursue adoption metrics to identify correlations between certain functions and ways to increase acceptance and, subsequent usage.

Additional ways to further this research include a follow up analysis to determine which apps are no longer active and provide reasoning on why that is.

## Conclusions

This global landscape analysis reveals how mobile apps are being used to manage the spread of COVID-19. The review of 224 COVID-19 mobile apps identified two main type of apps: Exposure Notification apps and non-Exposure Notification apps. Apps that do not serve to notify of exposure provide COVID related information and support. Research is needed to understand barriers and facilitators to adoption and how the variety of COVID-19 app features play into public acceptance and use.

## Data Availability

All data produced in the present study are available upon reasonable request to the authors

---

**Supplemental Table 1.**
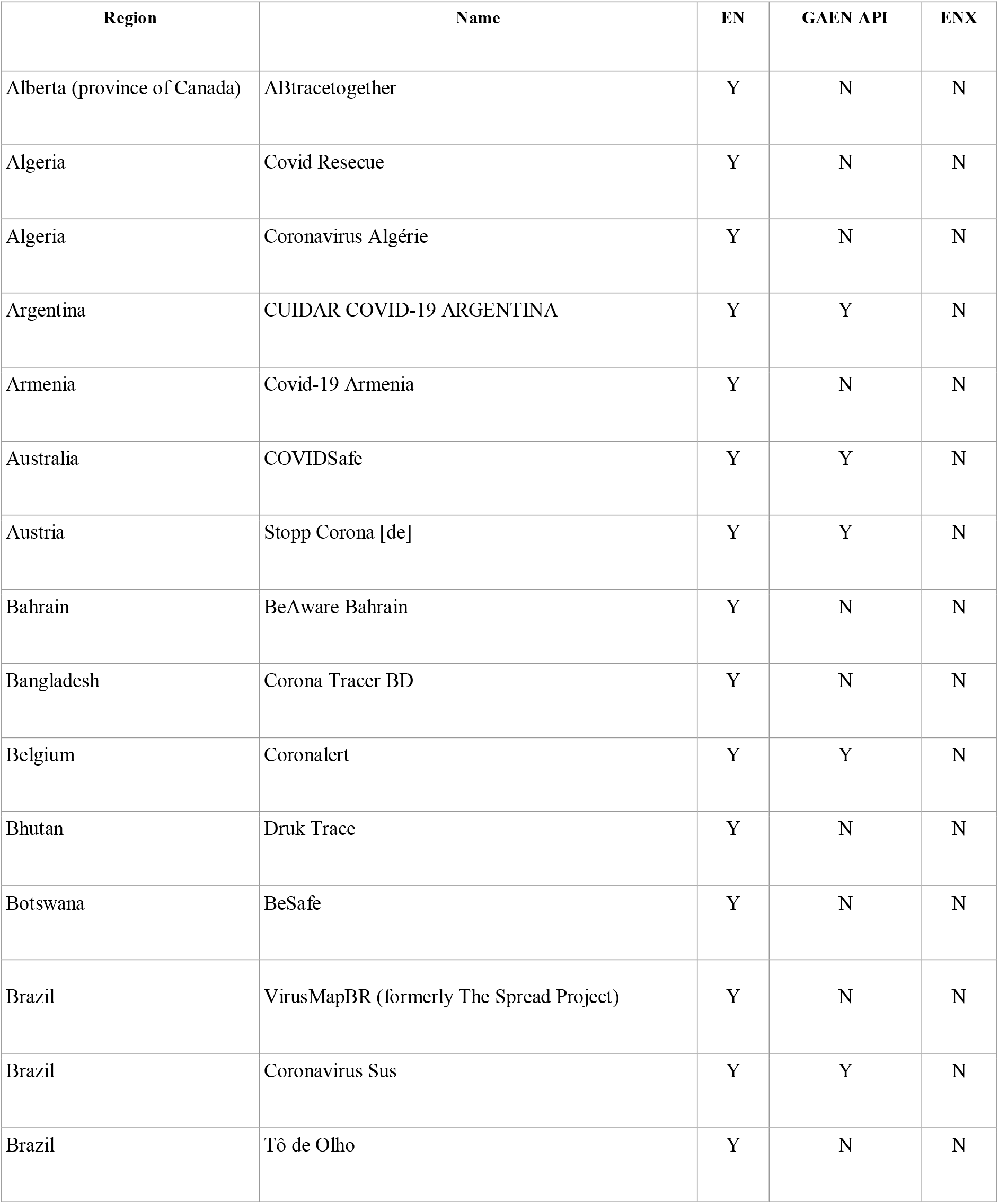
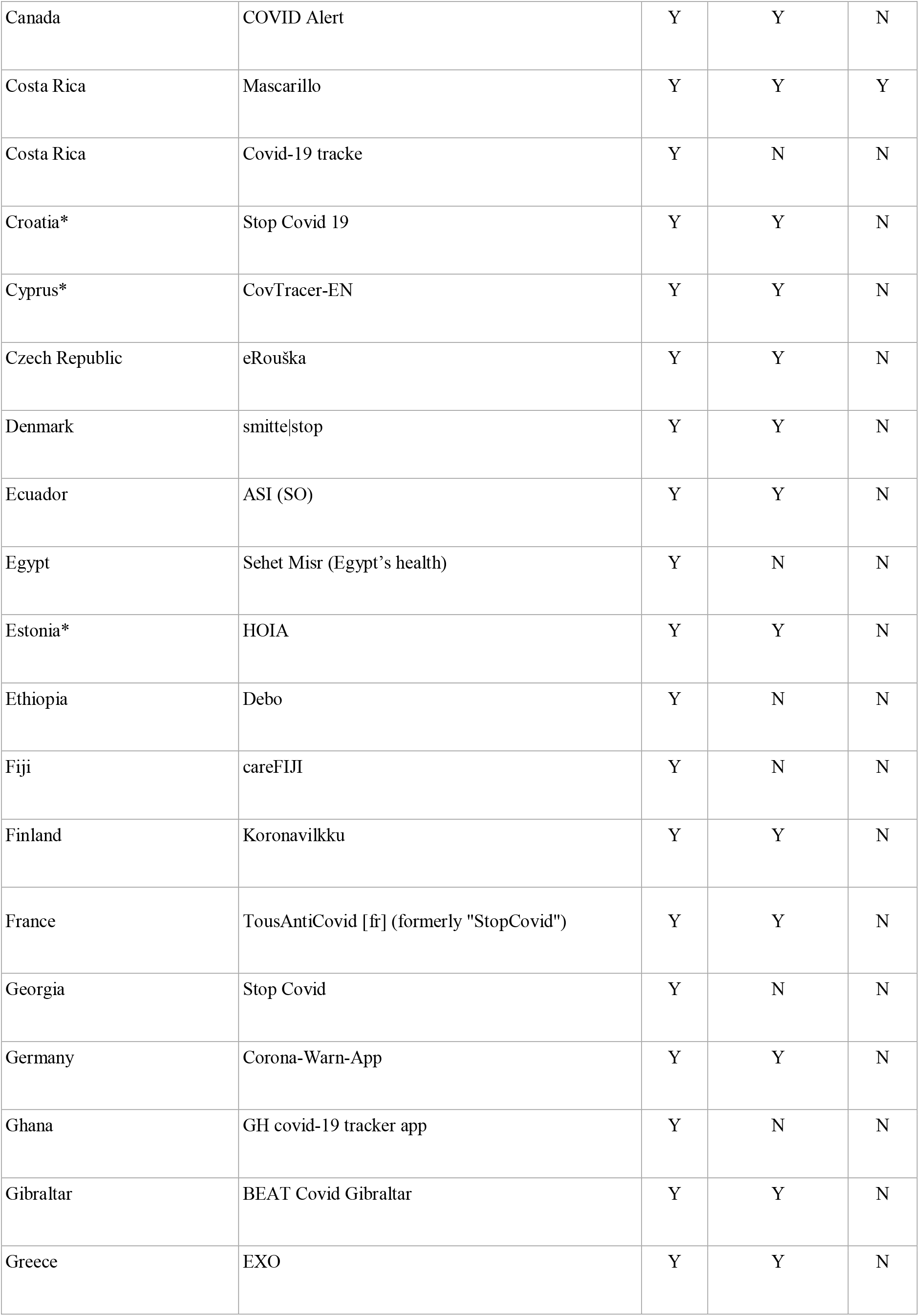
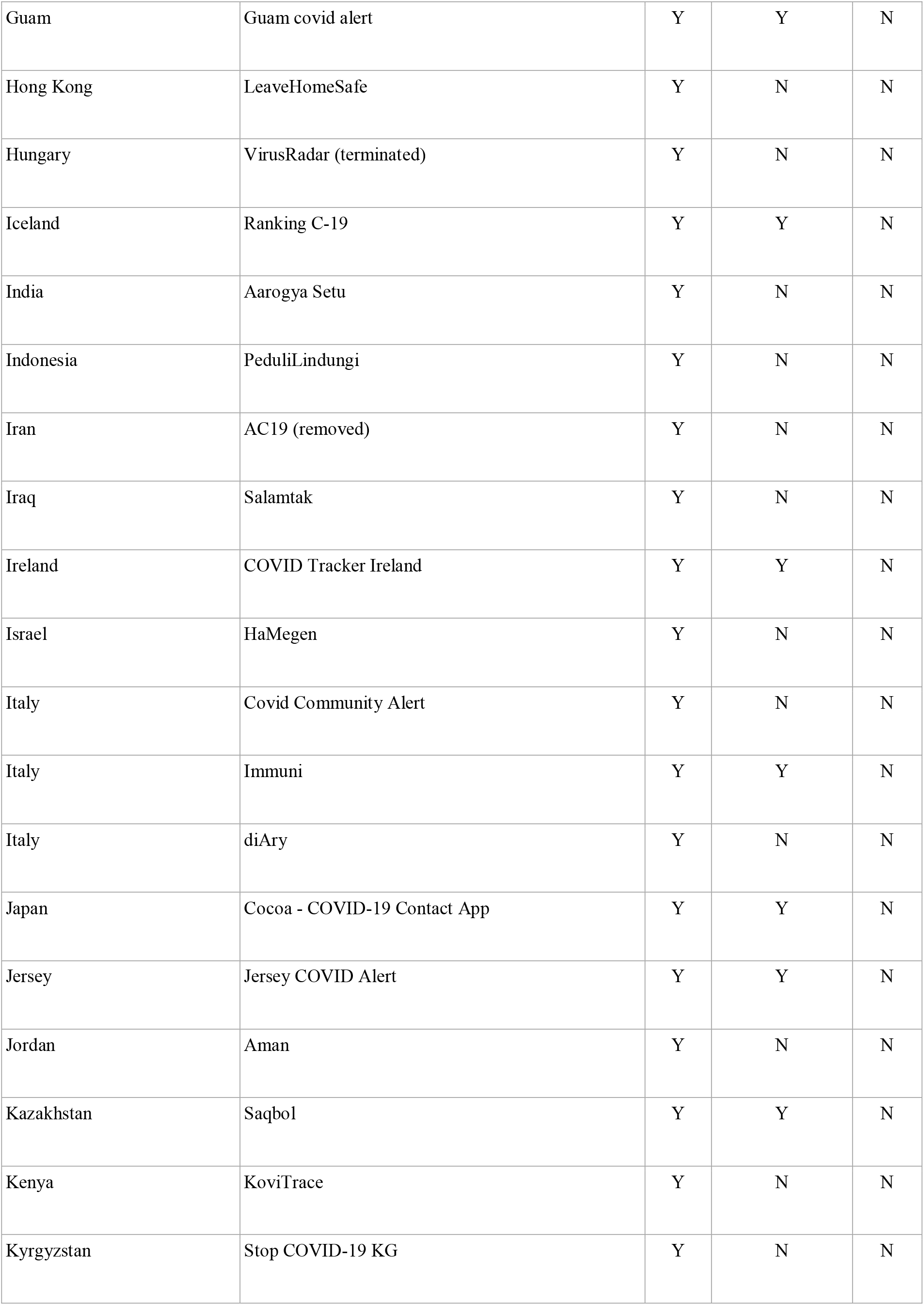
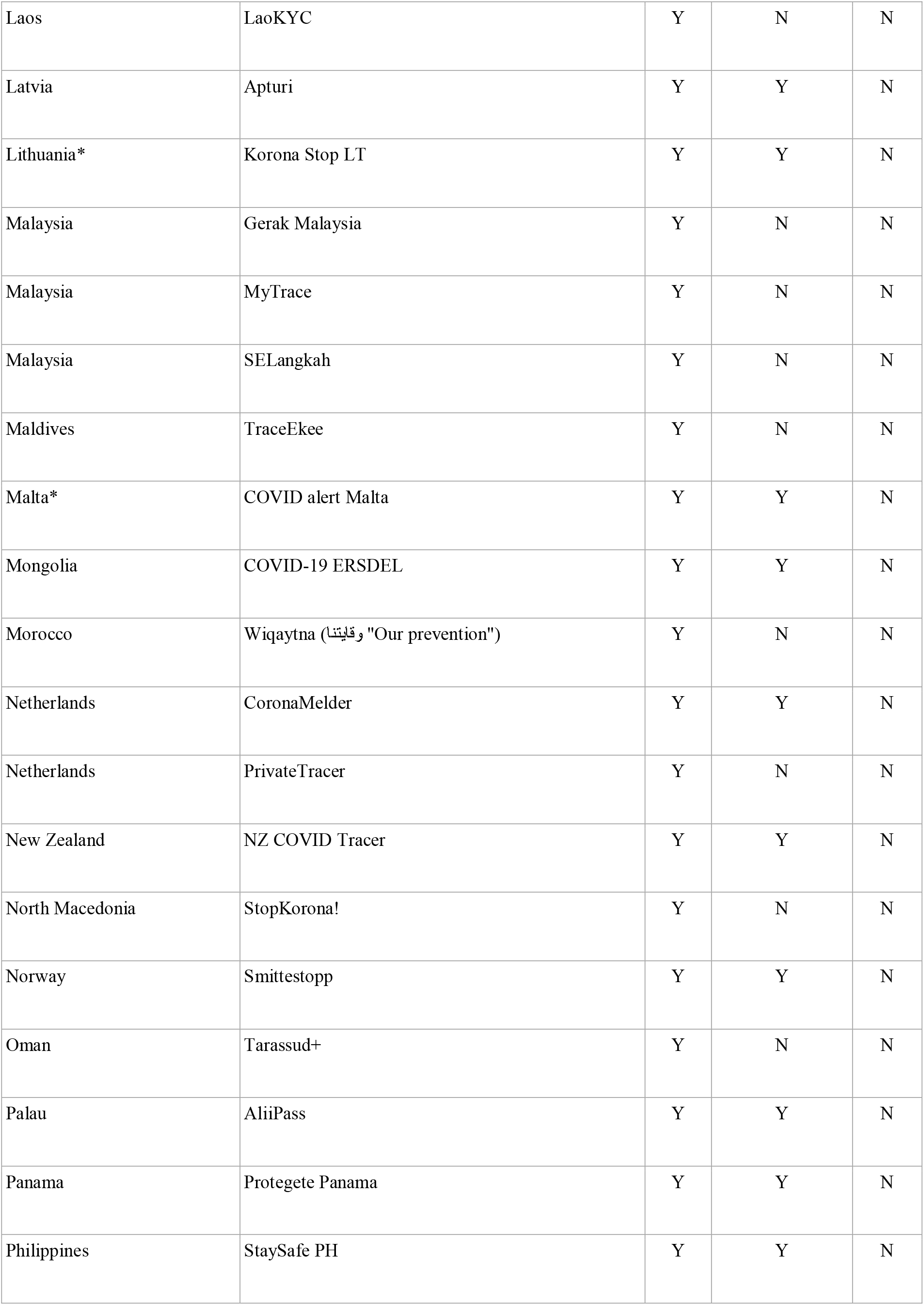
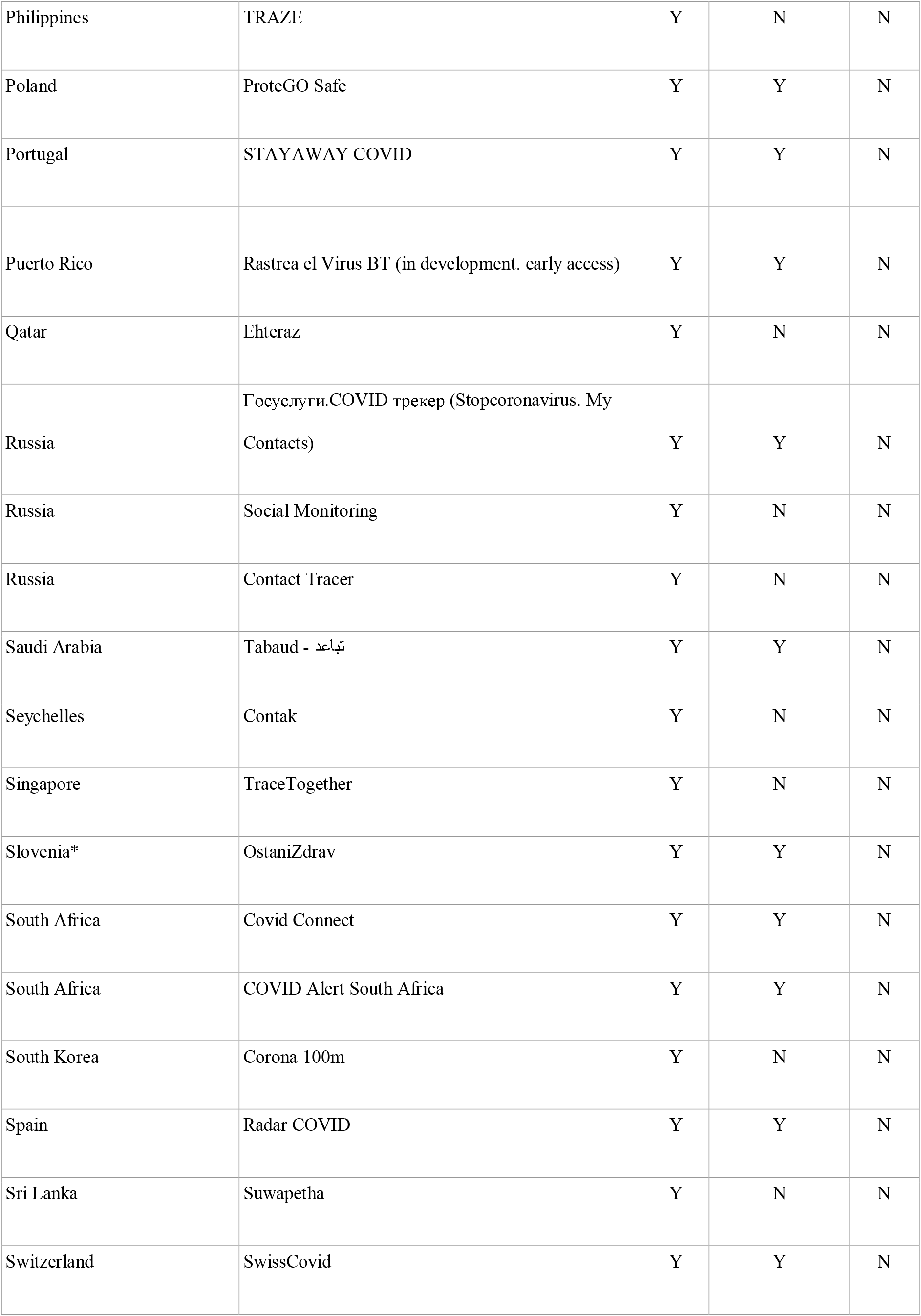
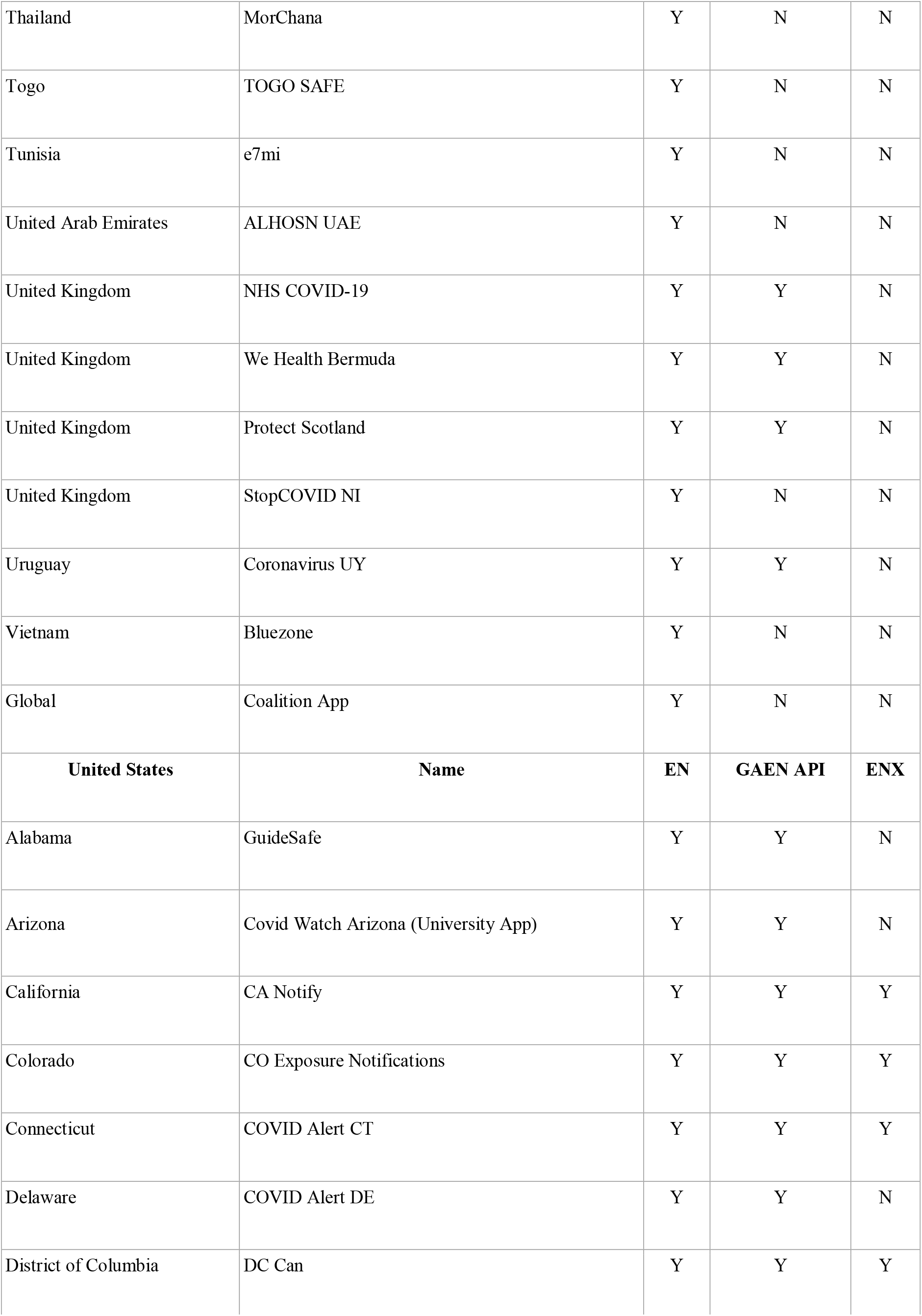
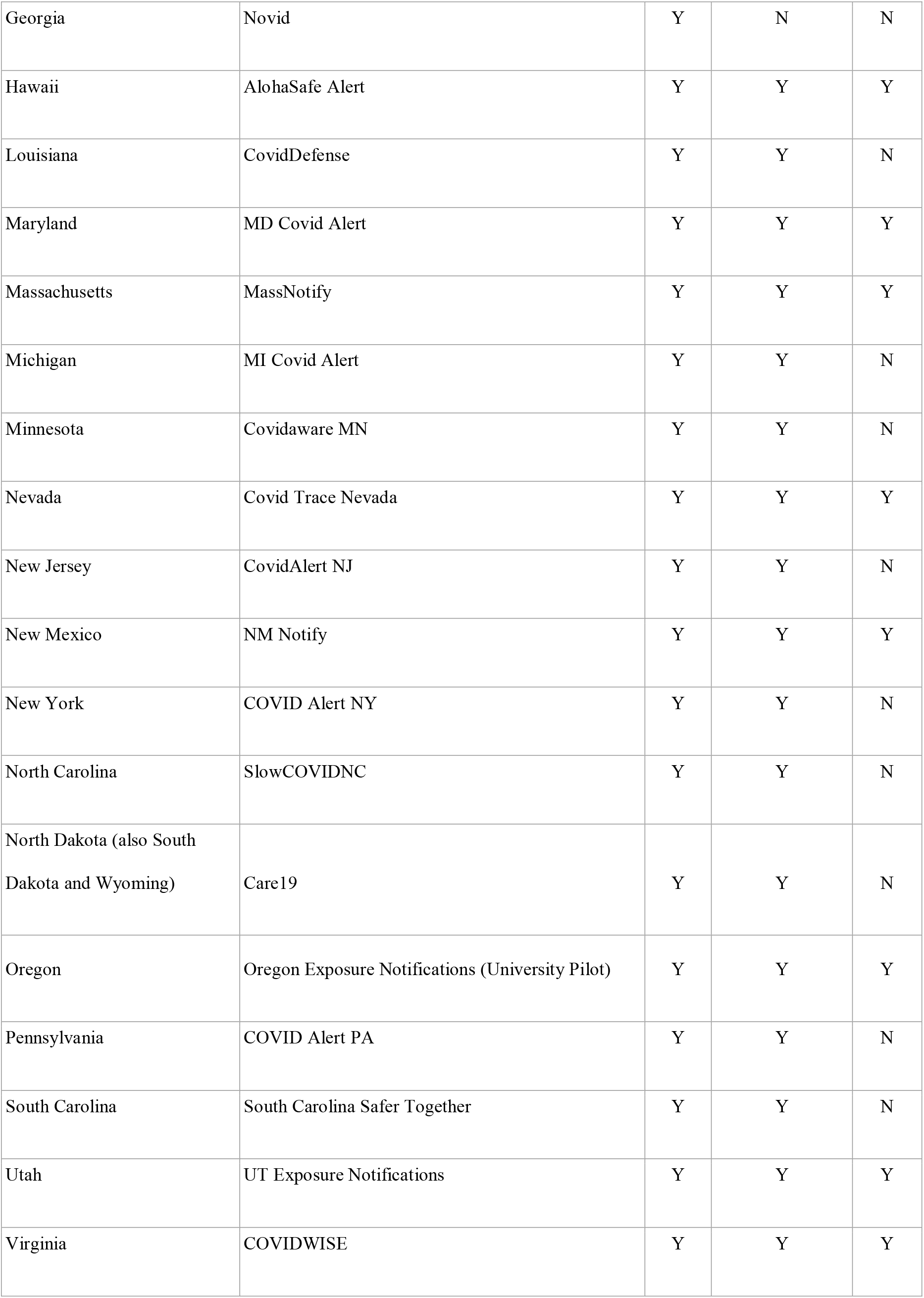
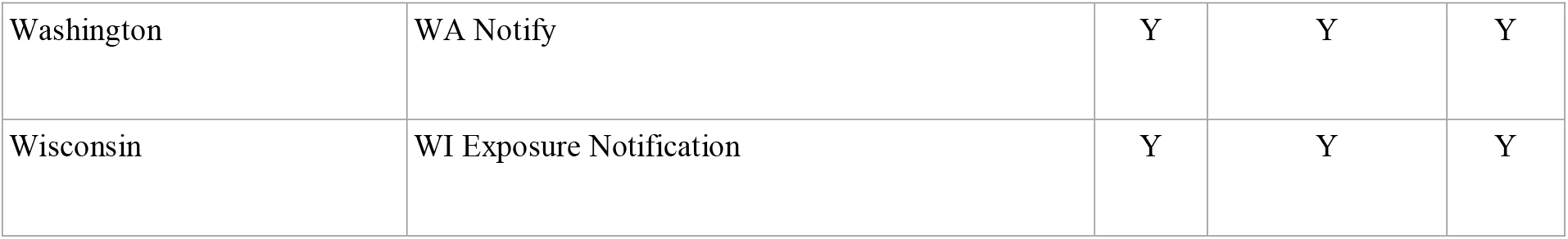
COVID-19 Exposure Notification Apps.

**Supplemental Table 2.**
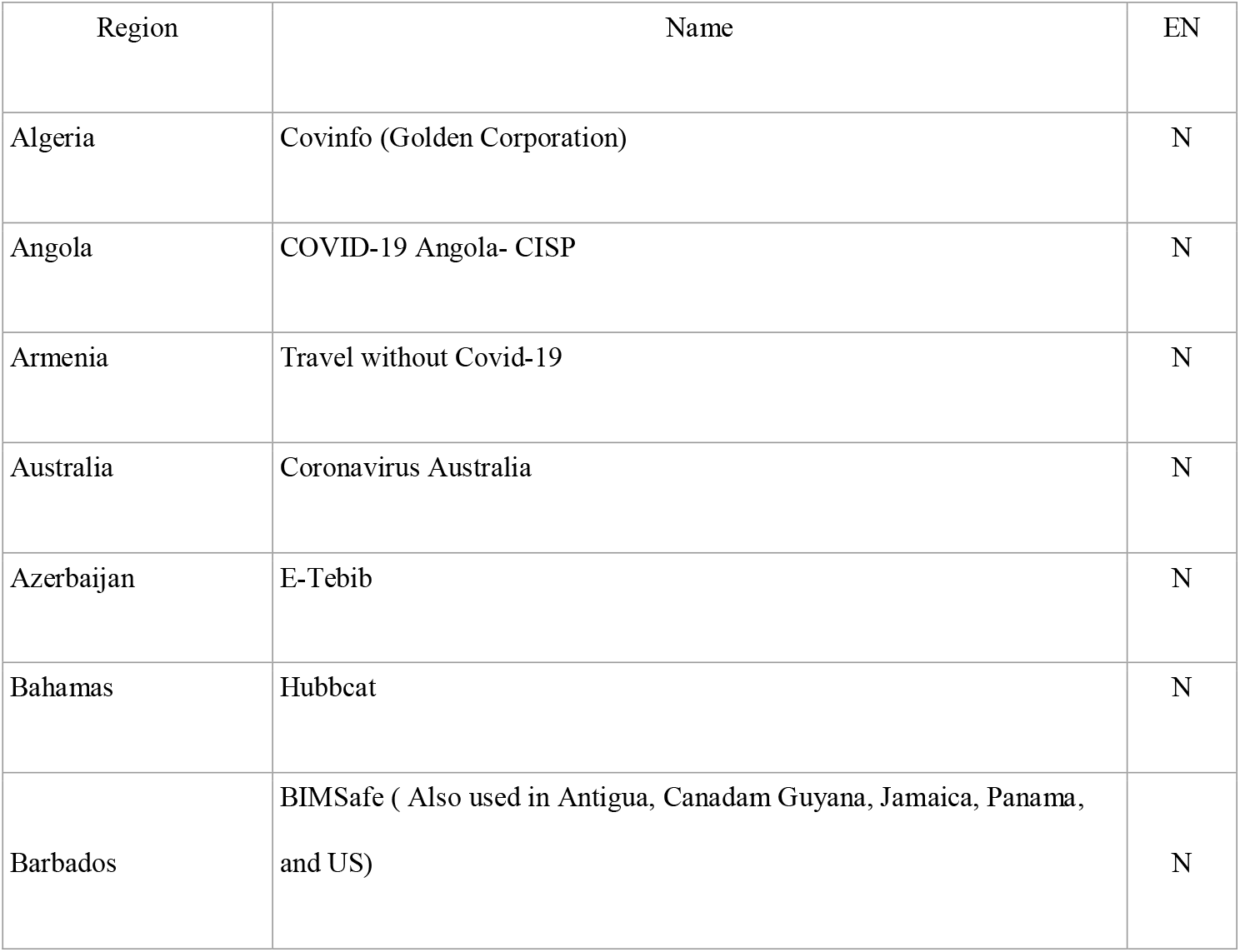
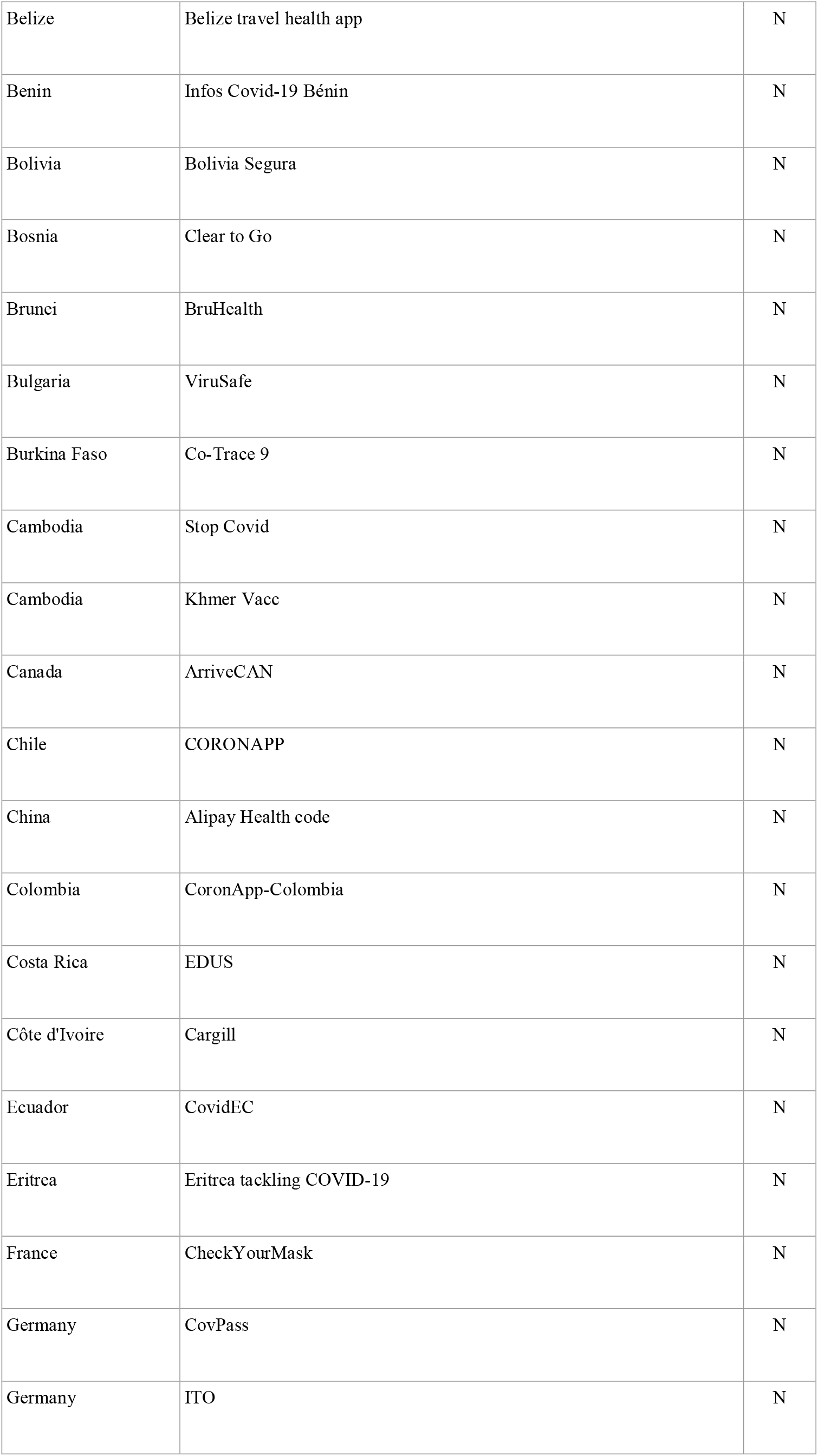
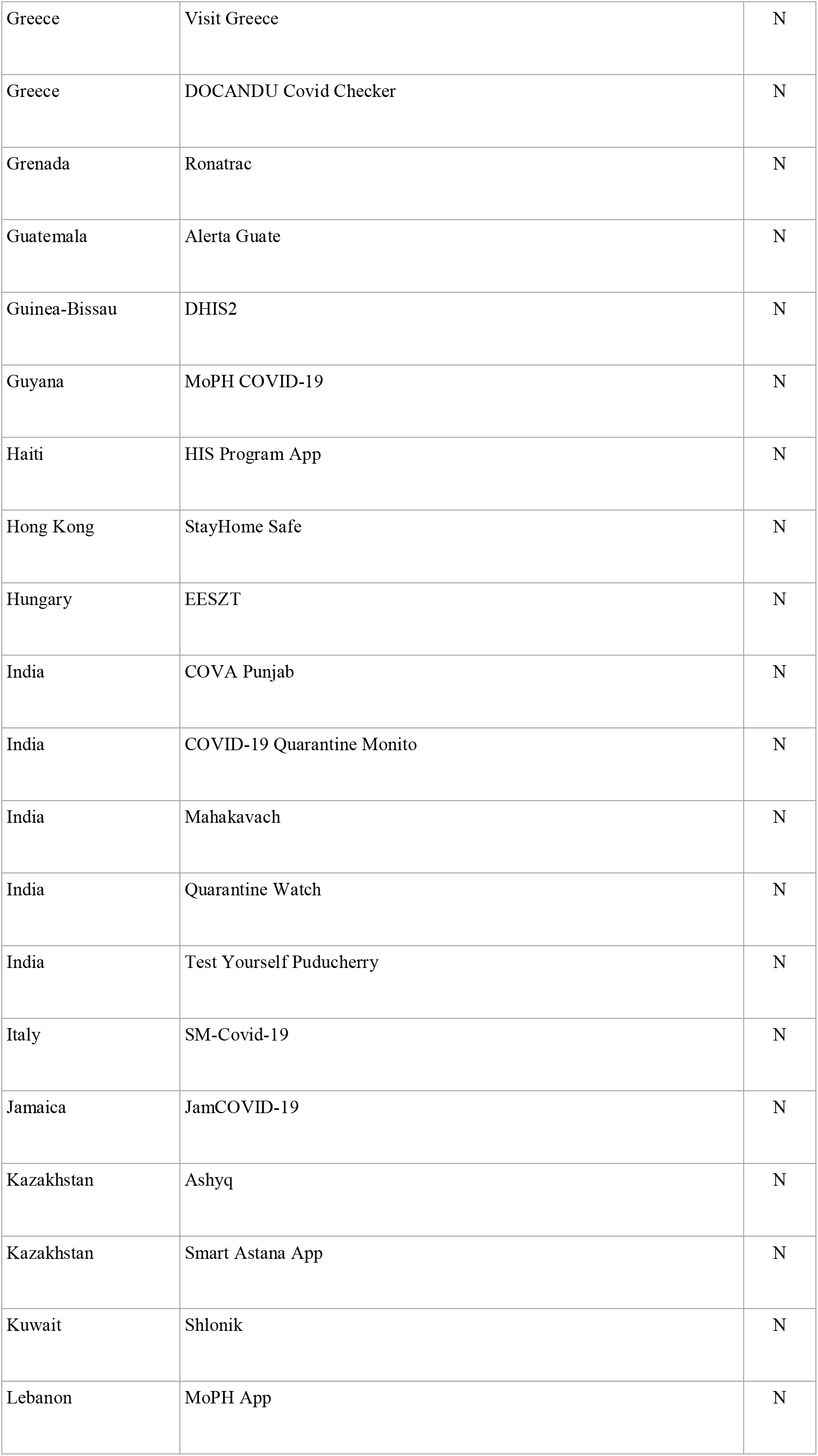
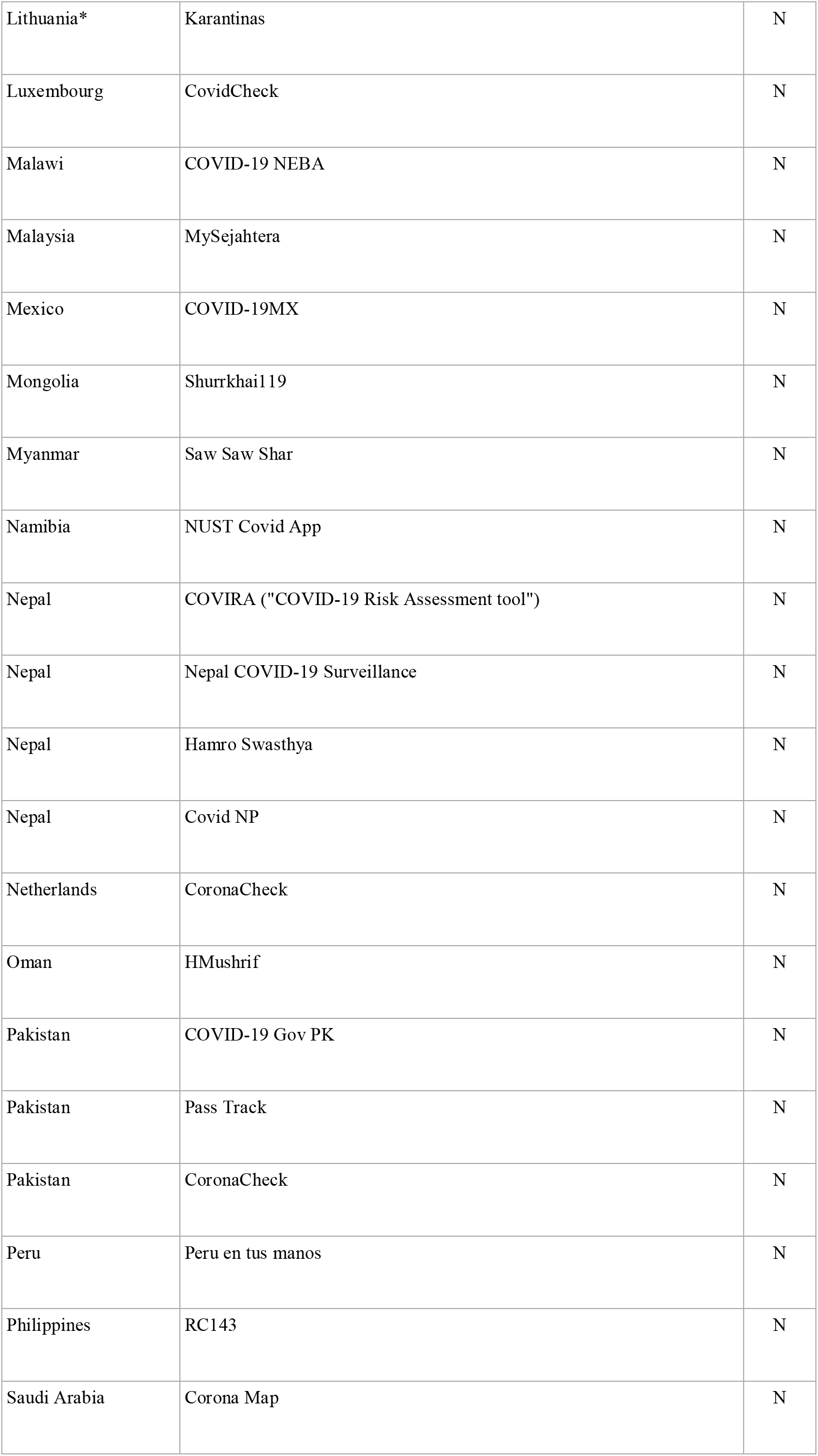
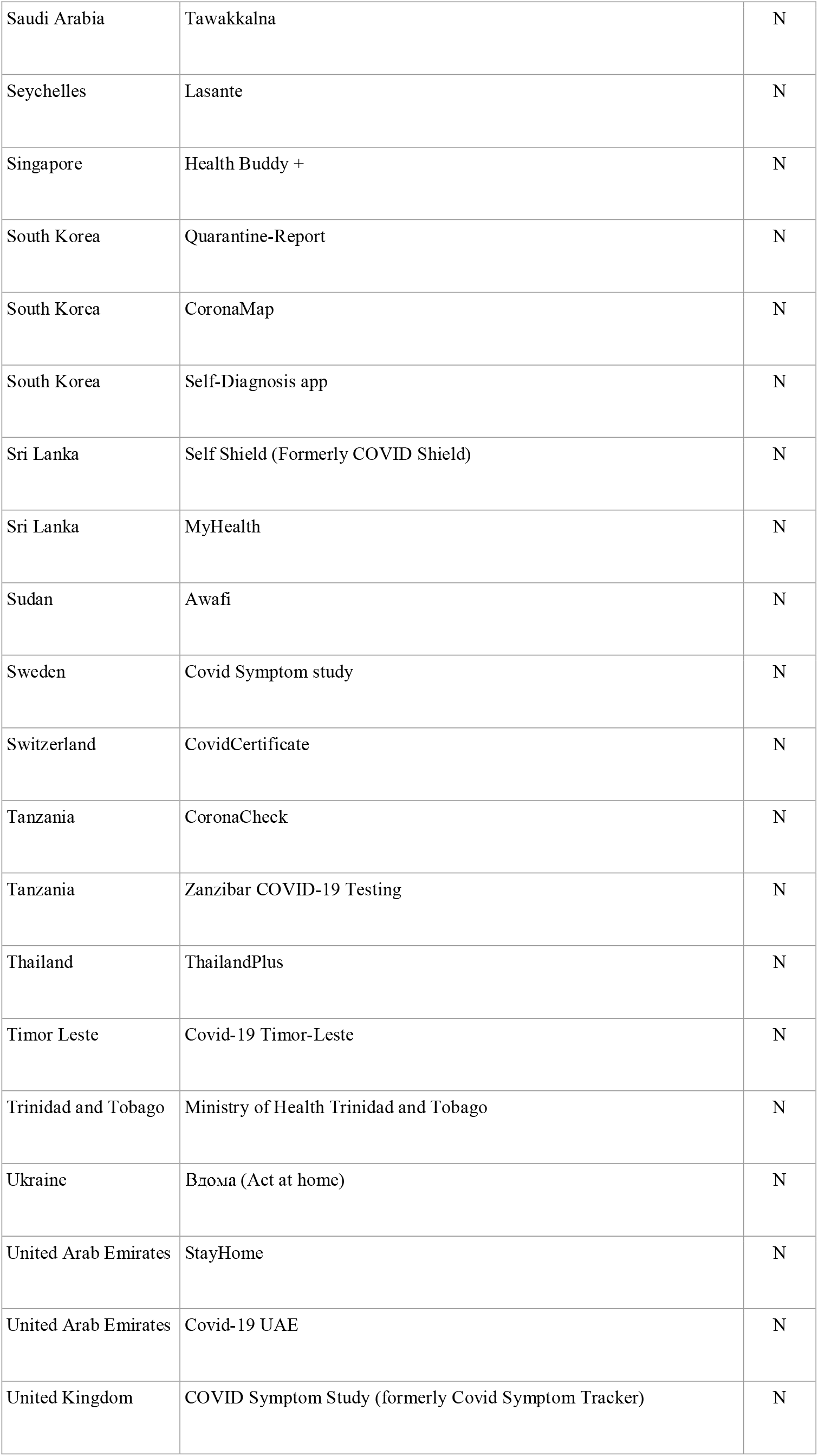
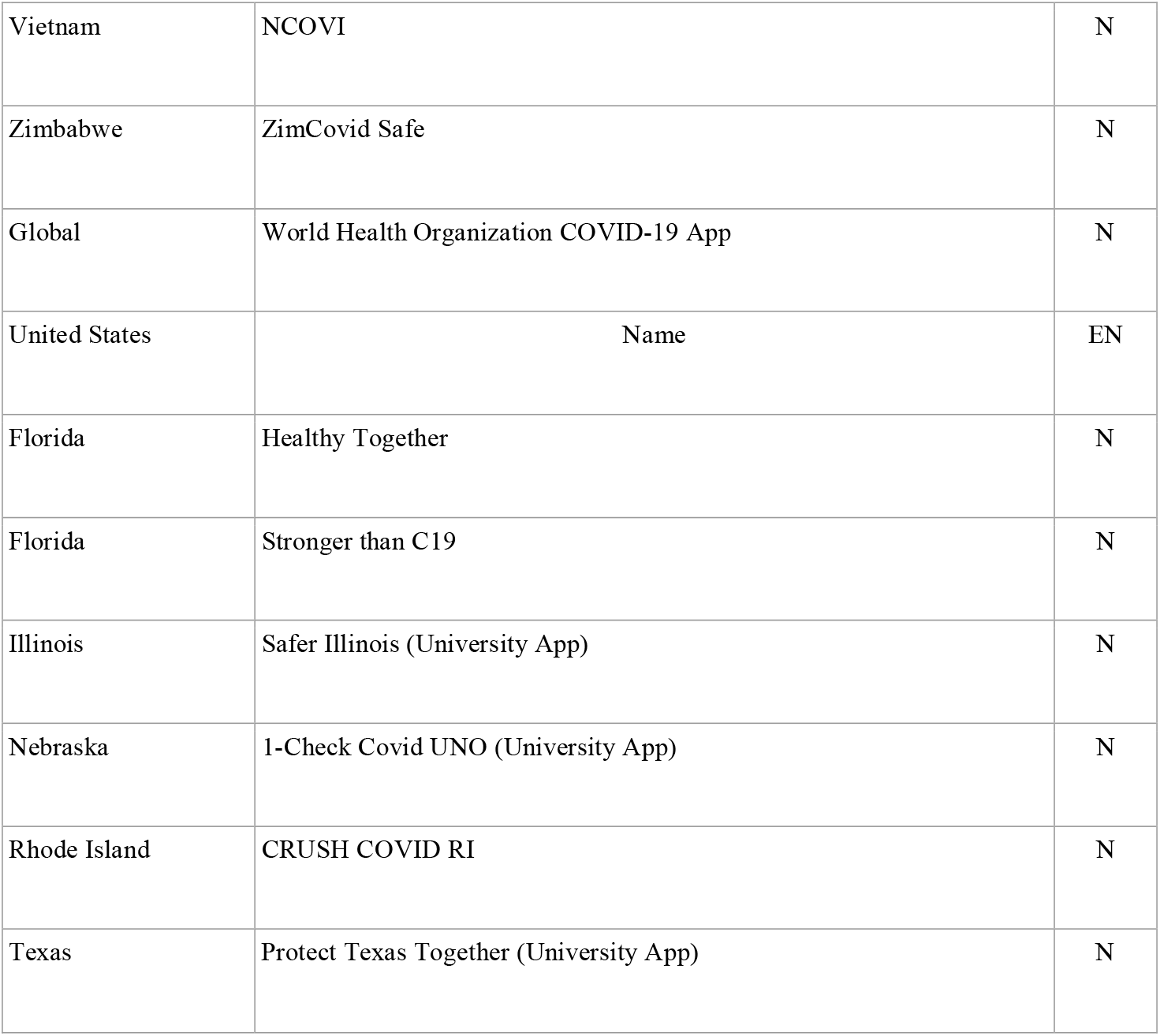
COVID-19 Non Exposure Notification Apps.

**Supplemental Table 3.**
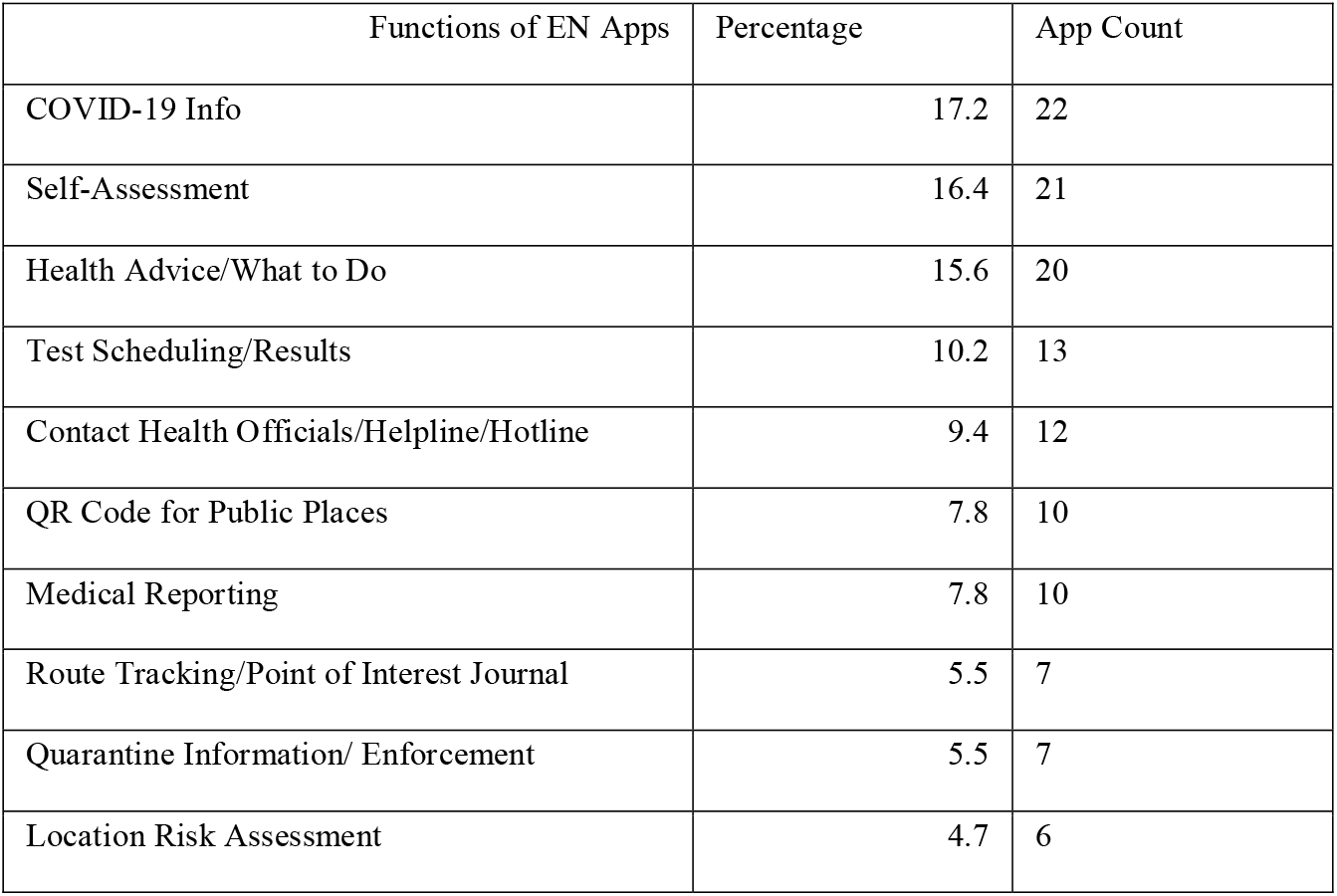
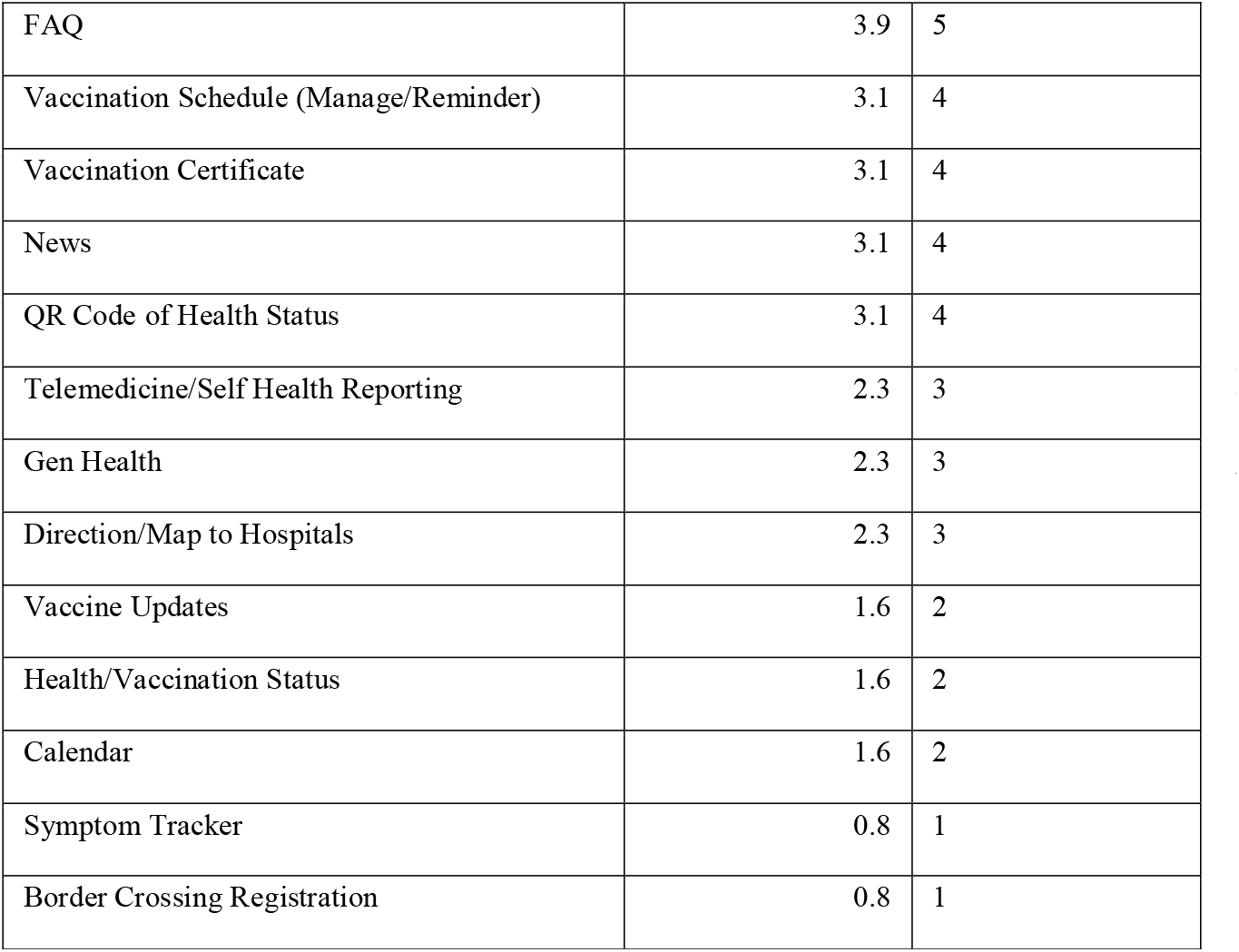
Features in EN Apps per Total Number of EN Apps.

**Table 4.**
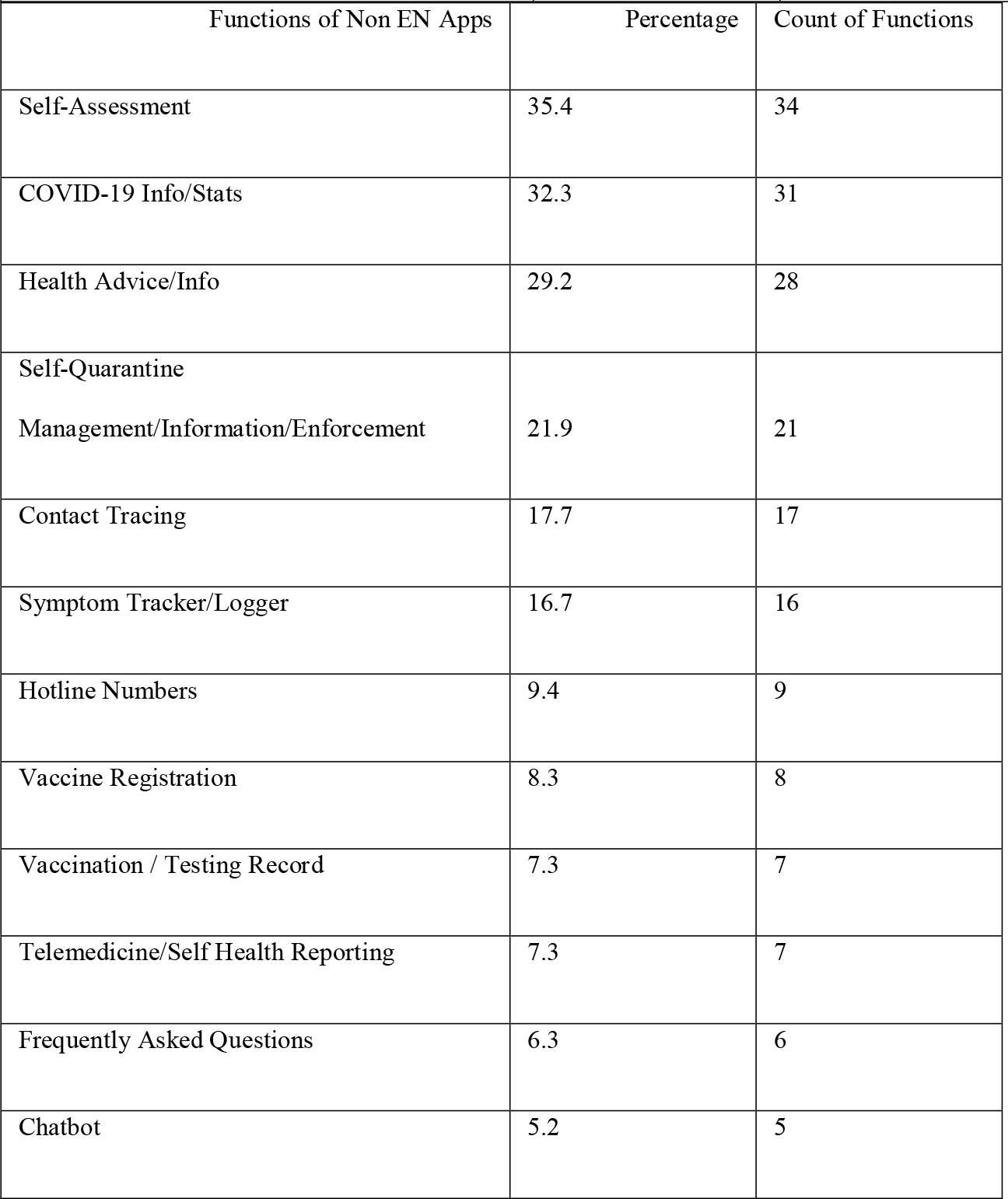
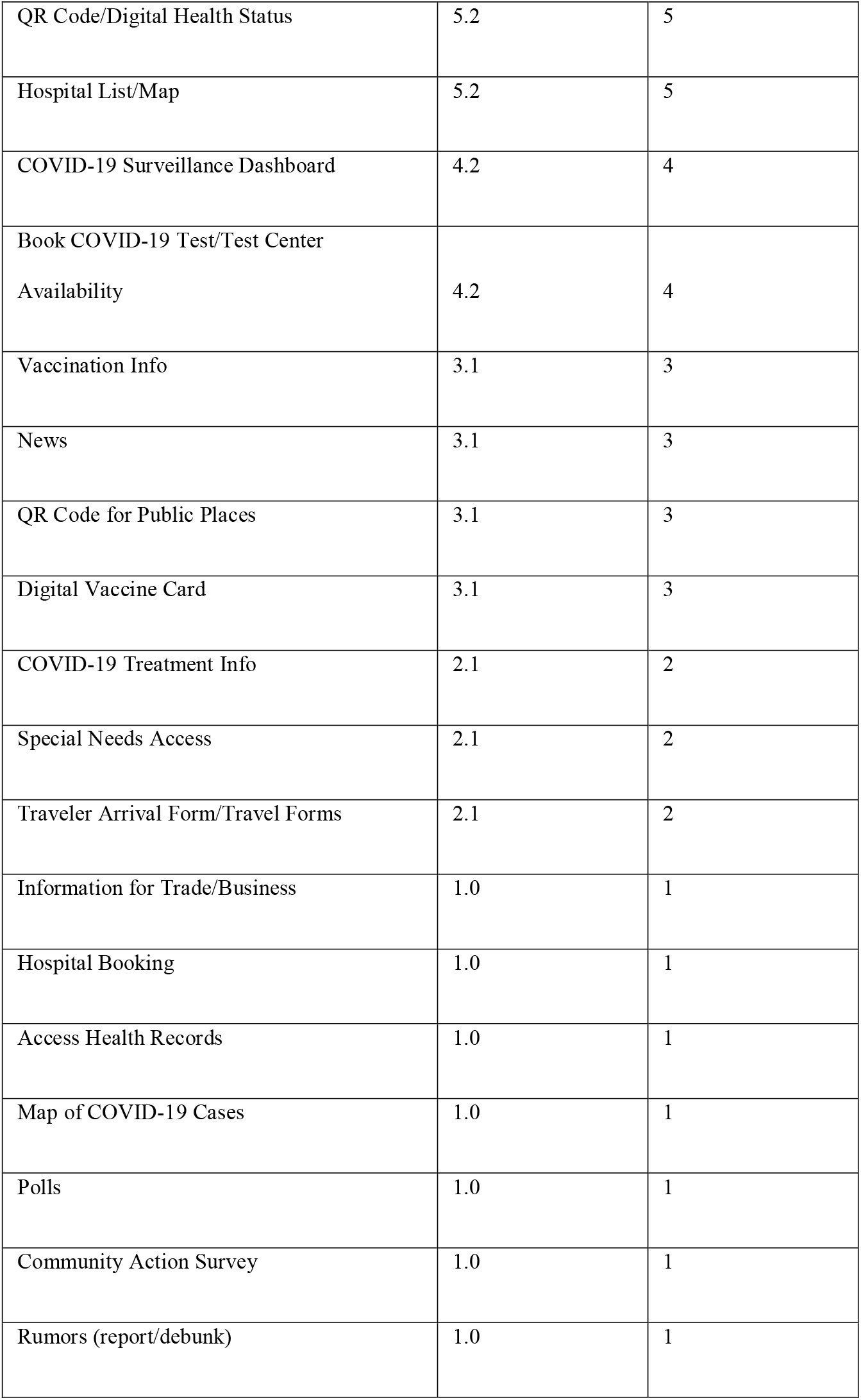
Non EN App Features per Total Number of non EN Apps.

**Table 5.** GAEN App Functions per Total Number of GAEN Apps.

| <u>Functions of EN Apps with GAEN API</u> | Percentage | Count |
| --- | --- | --- |
| COVID-19 Info | 17.3 | 13 |
| Self-Assessment | 16.0 | 12 |
| Health Advice/What to Do | 14.7 | 11 |
| Medical Reporting | 8.0 | 6 |
| Test Scheduling/Results | 8.0 | 6 |
| Contact Health Officials/Helpline/Hotline | 8.0 | 6 |
| FAQ | 5.3 | 4 |
| QR Code for Public Places | 2.7 | 2 |
| Vaccination Certificate | 2.7 | 2 |
| News | 2.7 | 2 |
| Location Risk Assessment | 2.7 | 2 |
| Direction/Map to Hospitals | 2.7 | 2 |
| Route Tracking/Point-of-Interest Journal | 2.7 | 2 |
| Calendar | 1.3 | 1 |
| General Health | 1.3 | 1 |
| Border Crossing Registration | 1.3 | 1 |
| Quarantine Information/ Enforcement | 1.3 | 1 |
| News | 1.3 | 1 |
| Symptom Tracker | 1.3 | 1 |
| Telemedicine | 1.3 | 1 |
| Health Status/Vaccination Status | 1.3 | 1 |

**Table 6.** Functions in non GAEN EN Apps per Total Number of non GAEN EN Apps.

| Functions of EN Apps with non GAEN API | Percentages | Count |
| --- | --- | --- |
| COVID-19 Info | 17.0 | 9 |
| Health Advice/What to do | 17.0 | 9 |
| QR code for Public Places | 15.1 | 8 |
| Self-Assessment | 15.1 | 8 |
| Contact Health officials/Hotline/Hotline | 11.3 | 6 |
| Test Scheduling/Results | 11.3 | 6 |
| Quarantine Information/ Enforcement | 11.3 | 6 |
| Route Tracking/Point-of-Interest Journal | 9.4 | 5 |
| QR code of Health Status | 7.5 | 4 |
| Vaccination Schedule (Manage/Reminder) | 7.5 | 4 |
| Medical Reporting | 7.5 | 4 |
| Location risk assessment | 7.5 | 4 |
| General Health | 3.8 | 2 |
| Vaccination Certificate | 3.8 | 2 |
| Vaccine Updates | 3.8 | 2 |
| Telemedicine/Self Health Reporting | 3.8 | 2 |
| Calendar | 1.9 | 1 |
| FAQ | 1.9 | 1 |
| News | 1.9 | 1 |
| Direction/Map to Hospitals | 1.9 | 1 |
| Health status/Vaccination Status | 1.9 | 1 |

## Footnotes

1* The notation (2+) indicates that more than 2 of that app’s type have been deployed within the region. EN apps not using the GAEN API were documented as a “non GAEN” app. The notation (2+) indicates that more than 2 of that app’s type have been deployed within the region. EN apps not using the GAEN API were documented as a “non GAEN” app. The documentation “GAEN + non GAEN” means that the region had deployed both a GAEN app and a non GAEN app. ENX (GAEN) means that the region deployed an Exposure Notifications Express app, which also uses the GAEN API.

## Notes

### Competing Interest Statement

The authors have declared no competing interest.

### Funding Statement

This study did not receive any funding

